# Single-cell analysis of bone marrow CD8+ T cells in Myeloid Neoplasms predicts response to treatment with Azacitidine

**DOI:** 10.1101/2023.12.30.23300608

**Authors:** Athanasios Tasis, Nikos E. Papaioannou, Maria Grigoriou, Nikolaos Paschalidis, Katerina Loukogiannaki, Anastasia Filia, Kyriaki Katsiki, Eleftheria Lamprianidou, Vasileios Papadopoulos, Christina Maria Rimpa, Antonios Chatzigeorgiou, Ioannis P. Kourtzelis, Petroula Gerasimou, Ioannis Kyprianou, Paul Costeas, Panagiotis Liakopoulos, Konstantinos Liapis, Petros Kolovos, Triantafyllos Chavakis, Themis Alissafi, Ioannis Kotsianidis, Ioannis Mitroulis

## Abstract

CD8^+^ T cells are critical players in anti-tumor immunity. In higher-risk myelodysplastic neoplasms (HR-MDS) and acute myeloid leukemia (AML), CD8^+^ T cells exhibit altered functionality, however whether this affects disease course is poorly understood. Herein, we aimed to identify immune cell signatures in the bone marrow (BM) associated with disease progression and treatment outcomes. In-depth immunophenotypic analysis utilizing mass and flow cytometry on 104 pre-treatment BM samples from patients with myeloid neoplasms, revealed an increased frequency of a CD57^+^CXCR3^+^ subset of CD8^+^ T cells in patients with HR-MDS and AML who failed AZA therapy. Furthermore, increased baseline frequency (>29%) of the CD57^+^CXCR3^+^CD8^+^ T cell subset correlated with poor overall survival. We further engaged scRNA-seq to assess the transcriptional profile of BM CD8^+^ T cells from treatment-naïve patients. We observed an increased abundance of cells within cytotoxic CD8^+^ T lymphocytes (CTL) cluster in secondary AML compared to HR-MDS. Additionally, response to AZA was positively associated with enrichment of IFN-mediated pathways, whereas enhanced TGF-β signaling signature in CTL clusters was observed in non-responders. Our results support that targeting of CD8^+^ T cells with inhibitors of TGF-β signaling in combination with AZA is a potential future therapeutic strategy in HR-MDS and AML.

## Introduction

Myelodysplastic neoplasms (MDS), chronic myelomonocytic leukemia (CMML) and acute myeloid leukemia (AML), are clonal disorders sharing common features in their pathobiology.^1^ While genetic alterations and epigenetic modifications are central in the pathobiology of myeloid neoplasms, the interaction between clonal and immune cells is also crucial,^2–4^ since compelling evidence indicate that the interplay between clonal cells and the bone marrow (BM) microenvironment plays a significant role in the development and progression of MDS and CMML.^5–7^ In particular, CD8^+^ T cells play a crucial role in the regulation of tumor microenvironment.^8^ Aberrant functionality of CD8^+^ T cells has been observed in patients with myeloid neoplasms compared to healthy individuals,^9–11^ rendering the former cell subset a major target for immunotherapeutic interventions.^12,13^

The standard of care for patients with higher-risk MDS (HR-MDS) and non-proliferating CMML, as well as patients with secondary AML, who are not eligible for intensive chemotherapy, is treatment with hypomethylating agents (HMA) like azacitidine (AZA).^14–16^ Additionally, AZA in combination with venetoclax is currently used in patients with previously untreated AML.^17^ However, not all patients exhibit a favorable response, and there is a significant risk of relapse.^18^ Additionally, molecular predictors of response to HMA and the precise mechanism of action of this drug are not well-defined. The exact genetic and cellular processes through which HMA exerts its effects are still being studied,^19^ whereas there is no therapeutic approach to overcome resistance to this treatment.^20^. Several lines of evidence suggest that AZA promotes the cellular and cytokine-mediated effector T-cell tumor lysis.^21,22^ Nevertheless, the exact role of CD8^+^ cells in disease progression and response to HMA in myeloid neoplasms remains to be defined.

The aim of the study was to comprehensively investigate the immune cell compartment in the BM of patients with myeloid neoplasms in various disease stages, to identify immune cell populations or molecular pathways that could predict response to treatment with HMAs and/or serve as targets for immunotherapies. For this reason, we initially engaged a systematic approach to study the immune landscape of BM samples by utilizing mass cytometry (Cytometry by time of flight; CyTOF) for the immunophenotypic characterization of immune cell populations in the BM. Based on this analysis, we then focused on BM CD8^+^ T cells and performed single-cell RNA sequencing (scRNA-seq), to address the molecular signature in CD8^+^ T cell subsets that are linked to response to treatment with AZA.

## Results

### Immunophenotypic analysis of BM immune cells from patients with myeloid neoplasms

To study the immune cell compartment, we performed deep immunophenotyping using CyTOF in BM samples from patients with LR-MDS (n=12), HR-MDS (n=15), AML (n=16) and CMML (n=5), collected before treatment initiation. Multidimensional scaling analysis revealed a separation of samples derived from patients with MDS and AML compared to CMML based on dimension 2, enforcing the notion of a different pathobiological background between MDS and the latter (Fig. 1A). Untargeted cluster analysis resulted in the identification of 14 clusters of cells (Fig. 1B, C). We observed a significantly decreased frequency of cells in the cluster of CD4^+^ T cells (CD4 T1) and B cells in patients with CMML, with a respective increase in the frequency of monocytic cells of myeloid cluster 2, characterized by the expression of CD11c, CD14 and CD38 (Fig. 1D). As chemokine signatures and the expression patterns of chemokine receptors potentially have prognostic implications in MDS and AML,^23,24^ we further evaluated the expression of the chemokine receptors C-C chemokine receptor 4 (CCR4), CCR6, CCR7 and C-X-C Motif Chemokine Receptor 3 (CXCR3), CD161 and CD294 within the T cell clusters (Supplementary Fig. 1). We observed increased expression of CXCR3 on cells from patients with AML and CMML compared to MDS patients in the CD4 T2, CD8 T1 and CD8 T2 clusters (Fig. 1E).

**Fig. 1.**
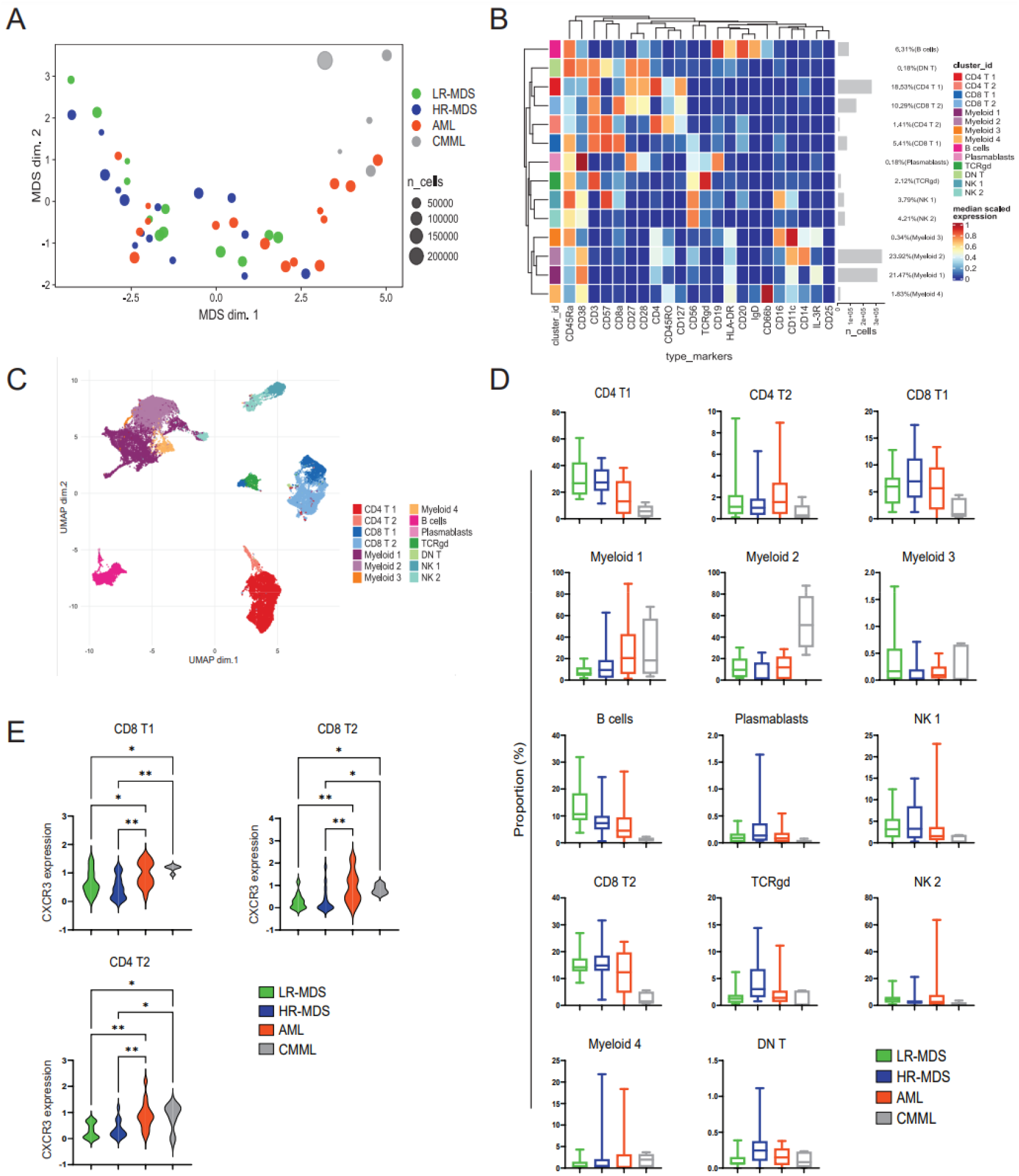
Untargeted analysis of CD45^+^ immune cells in patients with MDS, AML and CMML by CyTOF. (**A**) Multidimensional scale plot depicting the relationship between bone marrow (BM) samples of patients with LR-MDS (n=12), HR-MDS (n=15), AML (n=16) and CMML (n=5). (**B**) UMAP displaying the major immune cell clusters. (**C**) Heatmap showing the expression of the markers used for the characterization of each cell cluster. (**D**) Box charts displaying the frequency of each cell cluster. (Ε) Violin plots showing the expression level of CXCR3 in the CD8 T1, CD8 T2 and CD4 T2 clusters, respectively. Kruskal Wallis followed by “two-stage” Benjamini, Krieger & Yekutieli multiple comparison test was used in D. One-way ANOVA followed by “two-stage” Benjamini, Krieger & Yekutieli multiple comparison test was used in E. *p < 0.05, **p < 0.01.

Based on these findings, we then focused on CD4^+^ and CD8^+^ T cells and performed an untargeted cluster analysis on CD3^+^CD4^-^CD8^+^ (Fig. 2) and CD3^+^CD4^+^CD8^-^ T cells (Supplementary Fig. 2). We identified 10 clusters of CD8^+^ T cells (Fig. 2A). We noted a significantly decreased frequency of cells within cluster 1 in samples from patients with LR-MDS and HR-MDS compared to AML and CMML (Fig. 2B), a cluster that included terminal effector cells (CCR7^-^CD45RA^+^), expressing CD57 and CXCR3 (Fig. 2C). We also identified an additional cluster (cluster 2) of terminal effector cells (CCR7^-^CD45RA^+^), that did not express CD57 and CXCR3, and did not show any difference among the groups (Fig. 2B, C). Additionally, the expression of CXCR3 in cells from cluster 1 was higher in patients with AML and CMML compared to patients with MDS (Fig. 2D). Similar analysis on CD4^+^ T cells did not reveal any difference in the frequency of the generated clusters (Supplementary Fig. 2).

**Fig. 2.**
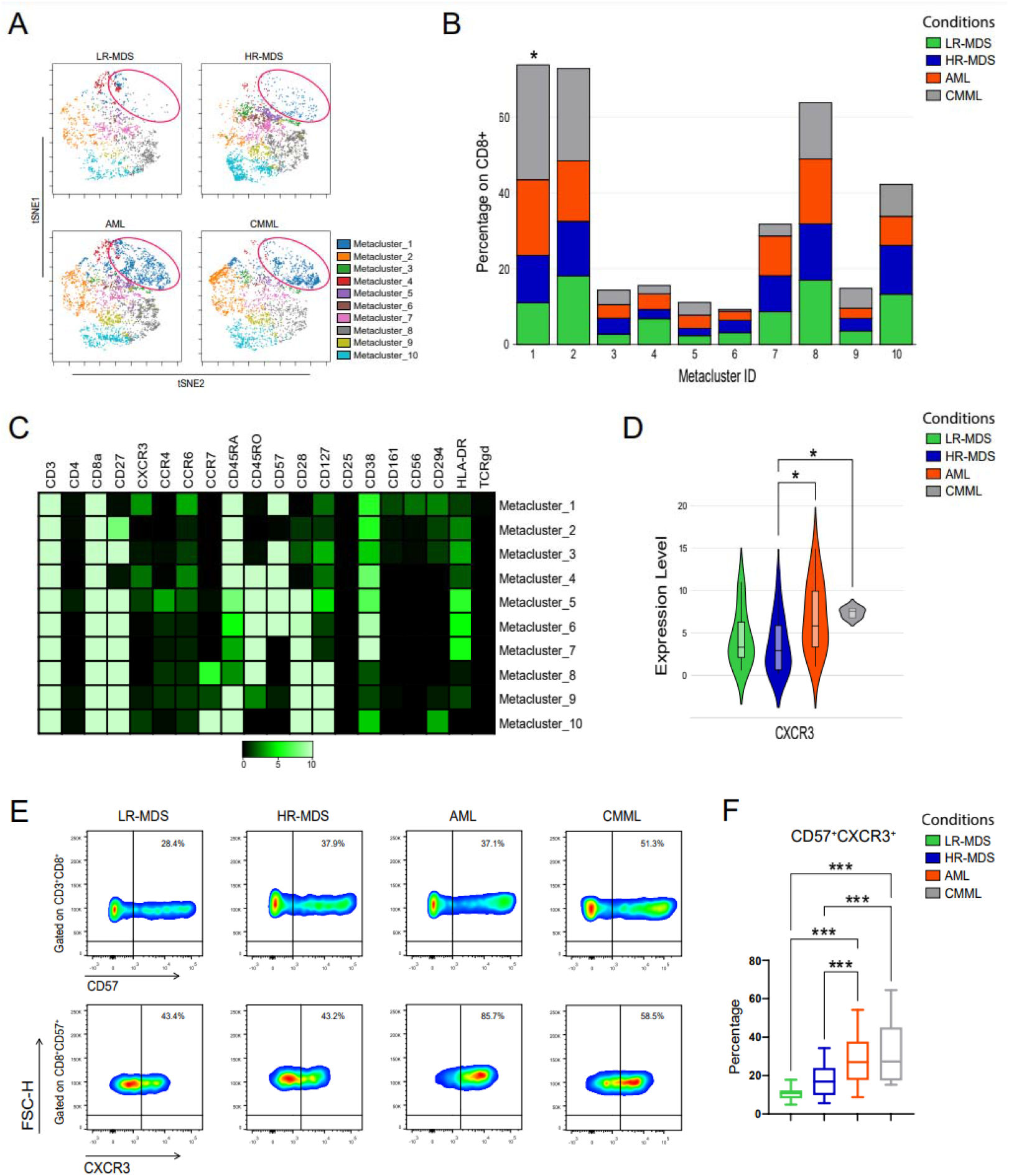
Identification of a CD8^+^ subpopulation (CD57^+^CXCR3^+^) which distinguishes MDS patients from AML and CMML patients. (**A**) Representative viSNE plots, derived from the FlowSOM analysis of BM CD8^+^ T cells from patients with LR-MDS (n=12), HR-MDS (n=15), AML (n=16) and CMML (n=5). (**B**) Bar plots displaying the proportion of the metaclusters between the groups, expressed as percentage within CD8^+^ T cells. (**C**) Heatmap depicting the expression level of the T-related markers between the metaclusters. (**D**) Violin plots showing the expression level of CXCR3 in metacluster 1. (**E**) Representative flow cytometry plots for the identification of the CD57^+^CXCR3^+^CD8^+^ T cell subpopulation in a cohort of patients with LR-MDS (n=7), HR-MDS (n=27), AML (n=20) and CMML (n=10). (**F**) Percentage of CD57^+^CXCR3^+^ cells within CD8^+^ T cells. Kruskal Wallis was used in B and D. One-way ANOVA followed by “two-stage” Benjamini, Krieger, & Yekutieli multiple comparison test was used in F. *p< 0.05, **p < 0.01, ***p < 0.001.

To further confirm the above findings, we engaged conventional flow cytometry in BM cells from an additional cohort of patients (n=64), focusing on the frequency of CD57^+^CXCR3^+^CD8^+^ T cells (Fig. 2E, F). We also analyzed with flow cytometry samples from eight patients that were also analyzed with CyTOF, to compare the two methods. We observed that the frequency of this cell population within CD8^+^ T cells was increased in patients with AML and CMML compared to LR- and HR-MDS (Fig. 2F), being in line with the findings from the unsupervised analysis of data derived from CyTOF. On the other hand, no difference was observed in the frequency of CD57^+^CXCR3^-^ CD8^+^ T cells (Supplementary Fig. 3).

Next, we characterized CD57^+^CXCR3^+^ CD8^+^ T cells compared to CD57^+^CXCR3^-^ CD8^+^ T cells, using the mass cytometry data. Increased expression of chemokine receptors CCR4 and CCR6 and decreased expression of the co-stimulatory molecules CD27 and CD28 was observed in CXCR3^+^ cells from patients with HR-MDS and AML (Supplementary Fig. 4). Additionally, flow cytometry analysis revealed decreased PD-1 expression in CXCR3^+^ cells compared to CXCR3^-^ cells (Supplementary Fig. 5).

### The frequency of CD57^+^CXCR3^+^CD8^+^ T cells is associated with the response to AZA

Dynamic changes of chemokine receptor expression on T cells may inform prognosis,^24^ whereas AZA may alter the BM chemokine profile in MDS patients. Having observed a gradual increase of the frequency of the CD57^+^CXCR3^+^CD8^+^ T cell subset from LR-MDS to AML, we next assessed whether the proportion of this cell population before treatment initiation with AZA was associated with response to treatment. Univariate analysis revealed that a decreased percentage of CD57^+^CXCR3^+^CD8^+^ T cells is associated with better response (Supplementary Table 1). Furthermore, multivariate analysis affirmed the independent predictive value of the baseline frequency of CD57^+^CXCR3^+^CD8^+^ T cells in relation to patient response (Supplementary Table 1). In line with this, utilizing the flow cytometry data, we observed that the baseline frequency of this cell subset was significantly increased in patients with HR-MDS and AML that did not respond to treatment, whereas no difference was observed in patients with CMML (Fig. 3A). Next, utilizing the CyTOF data, we performed unsupervised cluster analysis of CD3^+^CD4^-^CD8^+^ T cells in responders and non-responders to AZA patients with MDS and AML, which resulted in the identification of 6 cell clusters (Fig. 3B). We observed an increased frequency of cluster 3 in non-responders (Fig. 3B, C), a cluster of CCR7^-^CD45RA^+^ cells characterized by the expression of CD57 and CXCR3 (Fig. 3D). Conversely, we observed an increased frequency of cluster 6 in responders, characterized by naive/memory markers CD28, CD27, CCR7, CD127 (Fig. 3C, D), whereas there was no difference between responders and non-responders in the analysis of total CD3^+^CD8^-^CD4^+^ T cells (Supplementary Fig. 6).

**Fig. 3.**
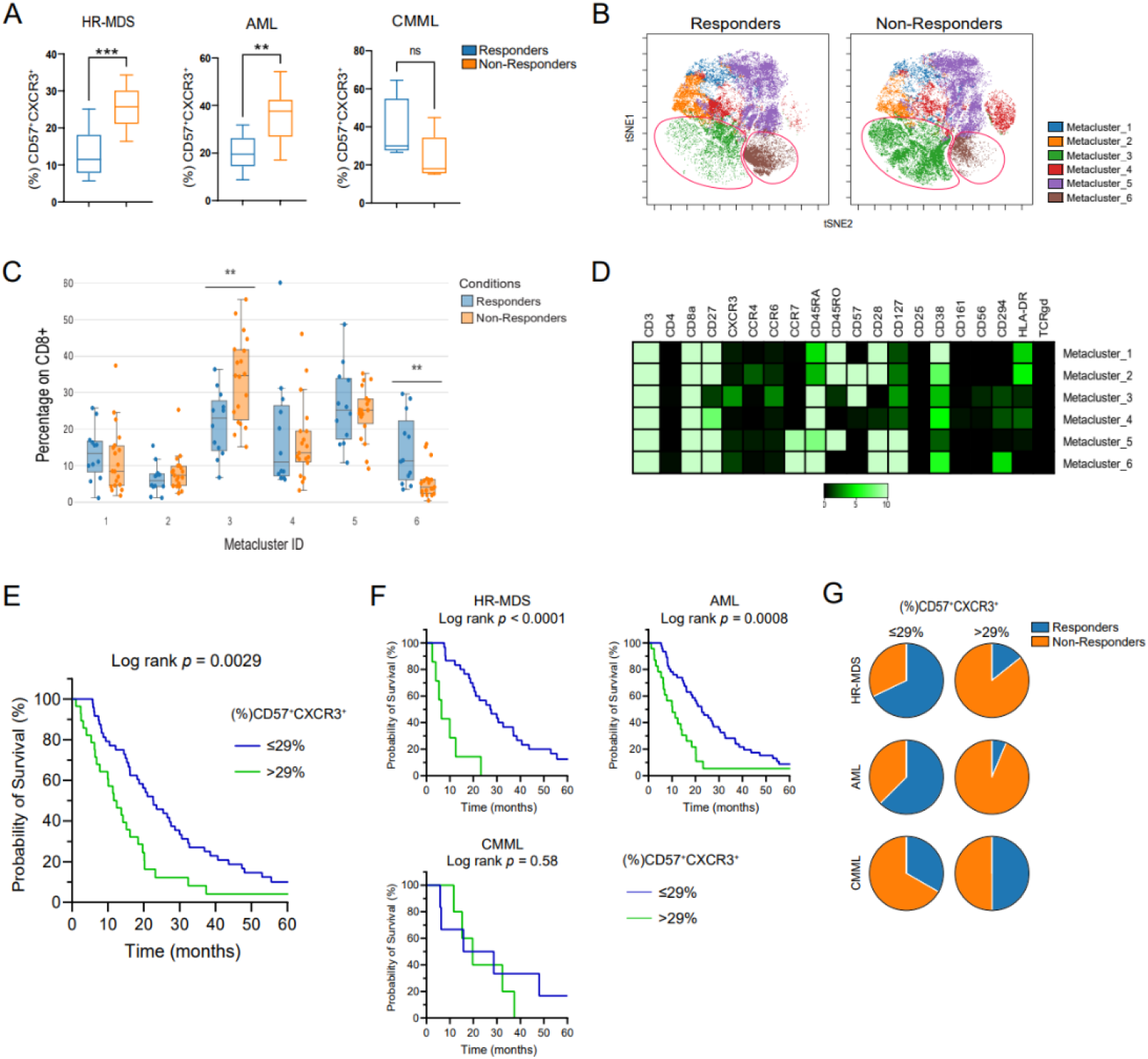
Association between the frequency of CD57^+^CXCR3^+^CD8^+^ T cells and outcome in patients with HR-MDS and AML under treatment with AZA. (**A**) Box plots displaying the percentage of the CD57^+^CXCR3^+^ cells within CD8^+^ T cells, assessed by flow cytometry in responders and non-responders (HR-MDS, n=12 Responders and 9 Non-Responders; AML, n=9 Responders and 11 Non-Responders; CMML, n=5 Responders and 5 Non-Responders). (**B**) After stratification of HR-MDS and AML patients to responders (n=12) and non-responders (n=19), FlowSOM analysis was performed on BM CD8^+^ T cells, which generated 6 metaclusters that are projected onto the viSNE plots. Representative viSNE plots (one for each group) are shown. (**C**) Heatmap depicting the expression levels of all T-related markers (**D**) Box plots showing the proportion of all metaclusters, expressed as frequency within CD8^+^ T cells. (**E**) Kaplan Meier curves for overall survival (OS) in patients which received AZA treatment, with ≤29% (n=51) and >29% (n=26) CD57^+^CXCR3^+^ CD8^+^ T cells before treatment initiation. The survival curves were compared by Log-rank (Mantel-Cox) test and the p value is shown. Median OS of the ≤29% group was 20.98 months, while the median OS of the >29% group was 12.05 months. (**F**) Survival curves for each disease subgroup. Increased (%) CD57^+^CXCR3^+^ correlates significantly with worse survival in HR-MDS and AML patients, whereas no association is observed in CMML patients. (**G**) HR-MDS and AML patients with ≤29% CD57^+^CXCR3^+^ exhibited higher response rates. No association between the frequency of CD57^+^CXCR3^+^CD8^+^ T cells and response to therapy was observed in CMML patients. Unpaired Student’s t test was used in A. Mann-Whitney U test was used in D. *p < 0.05, **p < 0.01, ***p < 0.001.

We further evaluated whether the baseline frequency of the CD57^+^CXCR3^+^CD8^+^ T cell subset was associated with survival. To do so, optimal CD57^+^CXCR3^+^ cut-off was determined at >29% by transformation of scale variable to binary one through optimal scaling. We observed that patients with ≤29% CD57^+^CXCR3^+^CD8^+^ T cells had better overall survival (OS) (Fig. 3E). However, when patients with HR-MDS, AML and CMML were evaluated separately, we observed that this cut-off value could be applied to patients with HR-MDS and AML but not to patients with CMML (Fig. 3F). Thereafter, we aimed to assess the applicability of this cut-off in predicting response to treatment with AZA. Patients with HR-MDS and AML characterized by a baseline frequency of ≤29% CD57^+^CXCR3^+^CD8^+^ T cells demonstrated higher rates of response to AZA (Fig. 3G). In contrast, the percentage of CD57^+^CXCR3^+^CD8^+^ T cells was not associated with response to treatment in patients with CMML (Fig. 3G), once more illustrating the distinct immune background of the latter. Together, these findings suggest that an increased frequency of the CD57^+^CXCR3^+^CD8^+^ T cell subset is associated with treatment failure and worse survival in patients with MDS and AML treated with AZA.

### Association between mutational status and frequency of CD57^+^CXCR3^+^CD8^+^ T cells

We further investigated whether there is a connection between mutational burden, type of mutations, and the frequency of CD57^+^CXCR3^+^CD8^+^ T cells in patients with HR-MDS and AML. No difference was found in the frequency of this cell population when patients were categorized based on the oncogenic mutations (Supplementary Fig. 7A), whereas no single mutation was associated with an increased frequency (>29%) of the CD57^+^CXCR3^+^CD8^+^ T cell subset (Supplementary Fig. 7B). Furthermore, the number of oncogenic mutations was not significantly altered in patients with >29% CD57^+^CXCR3^+^CD8^+^ T cells compared to patients with a lower frequency (≤29%) (Supplementary Fig. 7C), and no specific group of oncogenic mutations was associated with the frequency of CD57^+^CXCR3^+^CD8^+^ T cells (Supplementary Fig. 7D).

### Single-cell transcriptomic landscape of CD8^+^ T cells in patients with HR-MDS and secondary AML

We next sought to assess the molecular signature of BM CD8^+^ T cells from patients with MDS and AML secondary to MDS, at the single cell level, to further identify molecular signatures in specific CD8^+^ T cell subpopulations associated with disease progression and clinical outcomes of AZA monotherapy. scRNA-seq was performed in sorted BM CD8^+^ T cells from patients with HR-MDS (n=4) and secondary AML (n=5) prior to AZA initiation (Supplementary Table 2). We obtained transcriptomes of 28,449 cells in total. Based on unsupervised clustering, cells were partitioned in 11 clusters (Fig. 4A), which were characterized according to the gene expression of markers associated with T cell phenotype,^25^ including naive/memory markers (*CCR7*, *IL7R*, *SELL*, *CD27*, *CD28*, *CD44*), cytotoxic markers (*GZMA*, *GZMB*, *GZMK*, *PRF1*, *CX3CR1*, *NKG7*, *HOPX*, *KLRG1)*, cell cycle genes (*MKI67*, *CCNB2)*, the transcription factors (TFs) *LEF1*, *EOMES* and *TCF7*, the cytokine *IFNG*, and the cell surface markers *ITGA1*, *CD69* and *CCR6* (Fig. 4B, C). We identified clusters of cells characterized by the expression of *GZMK* (cluster 0), and *EOMES*, *KLRG1* and *GZMK* (cluster 1), previously characterized as pre-dysfunctional cells in studies from patients with solid tumors.^25,26^ We further identified two clusters of cytotoxic CD8^+^ T lymphocytes, clusters 2 (CTL_1) and 4 (CTL_2), based on the expression of *GZMA*, *GZMB* and *PRF1.*^27^ Clusters 1, 2 and 4 were the clusters with the highest frequency of cells expressing *B3GAT1*, the gene that encodes CD57 (Supplementary Fig. 8). We identified a cluster of memory-like cells, characterized by the expression of *IL7R* and low expression of *CCR7*, *SELL*, *CD27* (cluster 3), a cluster of naive-like cells, expressing *CCR7*, *SELL*, *CD27* and the TFs *TCF7* and *LEF1*^28^ (cluster 5), a cluster characterized by the expression of *CCR6* (cluster 6) and a cluster characterized by the highest expression of *CD44* (cluster 7) (Fig. 4B, C). Cluster 8 included cells expressing *ITGA1* and *ITGAE* (Fig. 4D), previously described as resident memory cells (cluster 8),^28–30^ and cluster 9 included cytotoxic cells expressing *IFNG*, *GZMK* and *NKG7*. Finally, a cluster of proliferating T cells (cluster 10) was identified (Fig. 4B, C). Top differentially expressed genes (DEGs) for each cell cluster are depicted in Fig. 4D. We further engaged a previously reported cytotoxic score^31^ to confirm the enhanced cytotoxicity of cells in clusters 2 (CTL_1), 4 (CTL_2) and 9 (*IFNG*) (Fig. 4E) and a cell cycle score^31^ to confirm the increased proliferation activity of cells in cluster 10 (Proliferative) (Fig. 4F). Finally, utilizing a previously reported dysfunctional/exhaustion score,^31^ we did not identify a specific cluster characterized by high expression of genes associated with exhaustion, such as *PDCD1*, *LAG3*, *HAVCR2*, *ENTPD1* or *CTLA4*^25^ (Supplementary Fig. 9A). Nevertheless, cluster 1 (*EOMES*) exhibited the highest dysfunctional score among the clusters with the highest cell abundancy (Supplementary Fig. 9B).

**Fig. 4.**
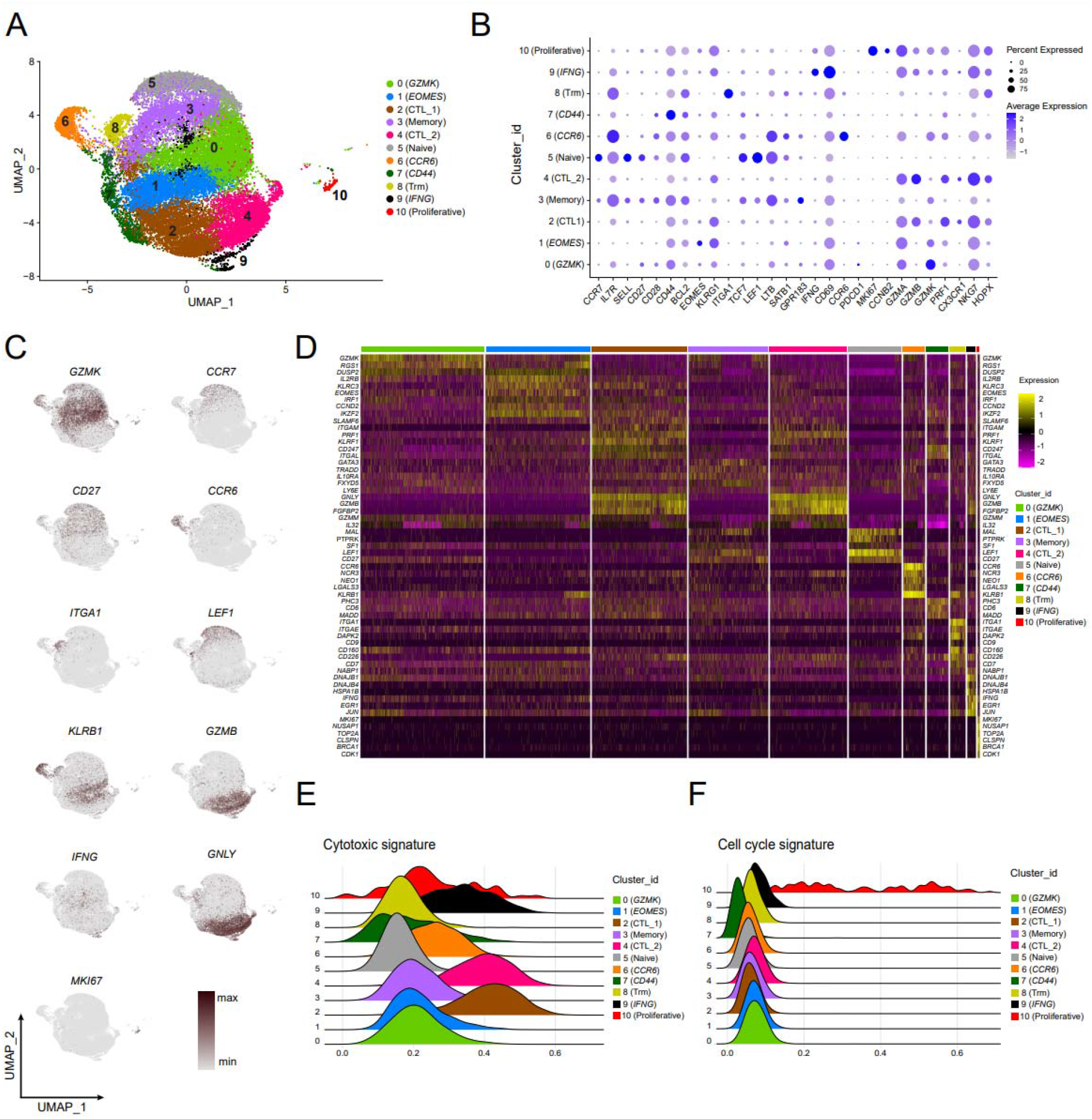
Profiling of BM-derived CD8^+^ T cells of HR-MDS and secondary AML patients with scRNA-seq. (**A**) Uniform Manifold Approximation and Projection (UMAP) of CD8^+^ T cells identified 11 clusters. A total of 28,449 CD8^+^ T cells were pooled from 4 HR-MDS (15,597 cells) and 5 secondary AML patients (12,852 cells). (**B**) Bubble plot depicting the average expression of genes used to characterize the clusters. (**C**) Expression of selected genes projected onto UMAPs. (**D**) Heatmap showing selected top differentially expressed genes for each cell cluster. (**E**) Ridgeline plots displaying the cytotoxic signature score for each cell cluster, as defined by the expression of key-related genes. (**F**) Ridgeline plots displaying the cell cycle signature score for each cell cluster. Differential expression data are reported in Data sheet 1.

We further studied whether there is a difference in the transcriptomic landscape of CD8^+^ T cells in HR-MDS compared to secondary AML (Fig. 5A). We observed that the frequency of cells in cluster 4 (CTL_2) was increased in patients with AML (Fig. 5B, C), a cluster showing high expression levels of *B3GAT1*, the gene that encodes the CD57 protein (Supplementary Fig. 8). Pathway analysis of the DEGs in cluster 4 (CTL_2) (Fig. 5D) revealed an overrepresentation of the T cell receptor pathway and anabolic pathways involved in DNA transcription, gene expression, mRNA processing and cell cycle in cells from patients with HR-MDS and IFN response pathways and oxidative phosphorylation (OXPHOS) pathway in cells from patients with AML (Fig. 5E). Of note, when we studied the exhaustion signature ^31^ between the two groups in cluster 1 (*EOMES*), a cluster characterized by the highest dysfunctional score among the clusters, we observed a significantly higher dysfunctional score in the HR-MDS group (Fig. 5F), which was associated with the enhanced expression of *CD7, FAM3C, TIGIT*, *TNFRSF9*, *DGKH*, *LYST*, *RAB27A*, *TNFRSF1B* (Fig. 7G) and *PDCD1, CD244*, *AKAP5*, *KIR2DL4* (Supplementary Fig. 10).

**Fig. 5.**
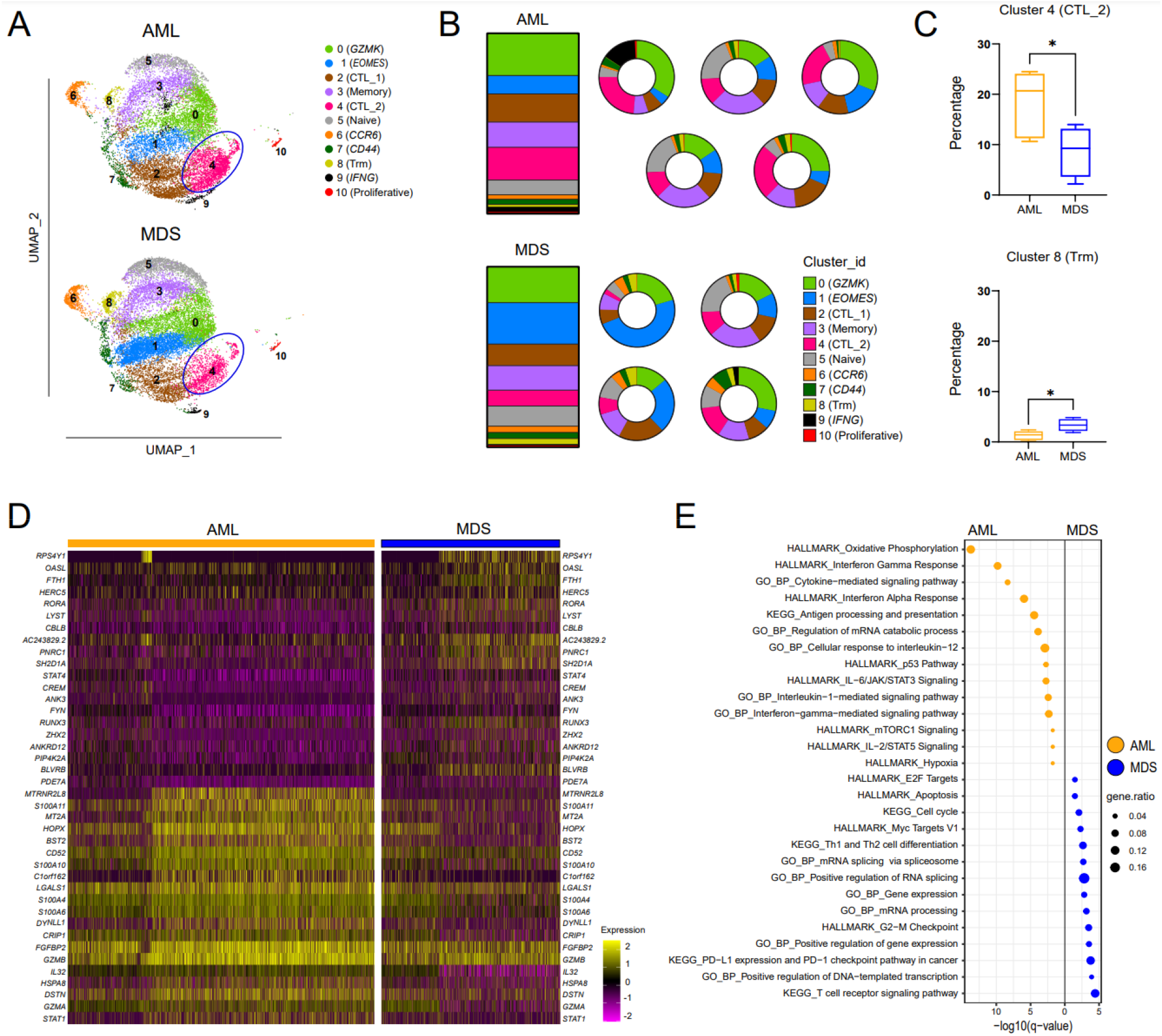
scRNAseq analysis of bone marrow-derived CD8^+^ T cells from patients with secondary AML and HR-MDS. (**A**) Comparison of separate UMAPs for secondary AML (12,852 cells) and HR-MDS (15,597 cells) patients. (**B**) Stacked bar chart showing the average distribution of clusters between the patient groups, and pie charts showcasing the distribution of clusters for each patient, individually. (**C**) Boxplot showing the percentage of clusters 4 (CTL_2) and 8 (Trm) expressed as (%) of total CD8^+^ T cells. (**D**) Heatmap displaying the top 20 differentially expressed genes of each disease group in cluster 4 (CTL_2). (**E**) Pathway enrichment analysis (MSigDB, KEGG and GO_BP) of cluster 4 (CTL_2) between HR-MDS and secondary AML patients. Positively enriched pathways in each group, with a q-value <0.05 (Benjamini-Hochberg correction), are shown. (**F**) Dysfunctional score for cluster 1 (*EOMES*). (**G**) Violin plots displaying the expression levels of the top differentially expressed genes involved in the dysfunctional score of cluster 1 (*EOMES*) between patients with HR-MDS and secondary AML. Differential expression data are reported in Data sheet 2.

### Single-cell transcriptomes of CD8^+^ T cells are associated with treatment outcome

We next sought to identify molecular signatures in CD8^+^ T cell subpopulations associated with response to AZA (Fig. 6A). We did not observe any statistically significant difference in the cluster abundance in patients that achieved complete remission (CR) after treatment with AZA (n=2 patients with HR-MDS, 7,571 cells; n=2 patients with secondary AML, 6,096 cells) and patients that failed (FAIL) to treatment (n=2 patients with HR-MDS, 8,026 cells; n=3 patients with secondary AML, 6,756 cells) (Fig. 6B), except from an increased abundancy of cluster 10 (Proliferative) in the CR group (Supplementary Fig. 11).

**Fig. 6.**
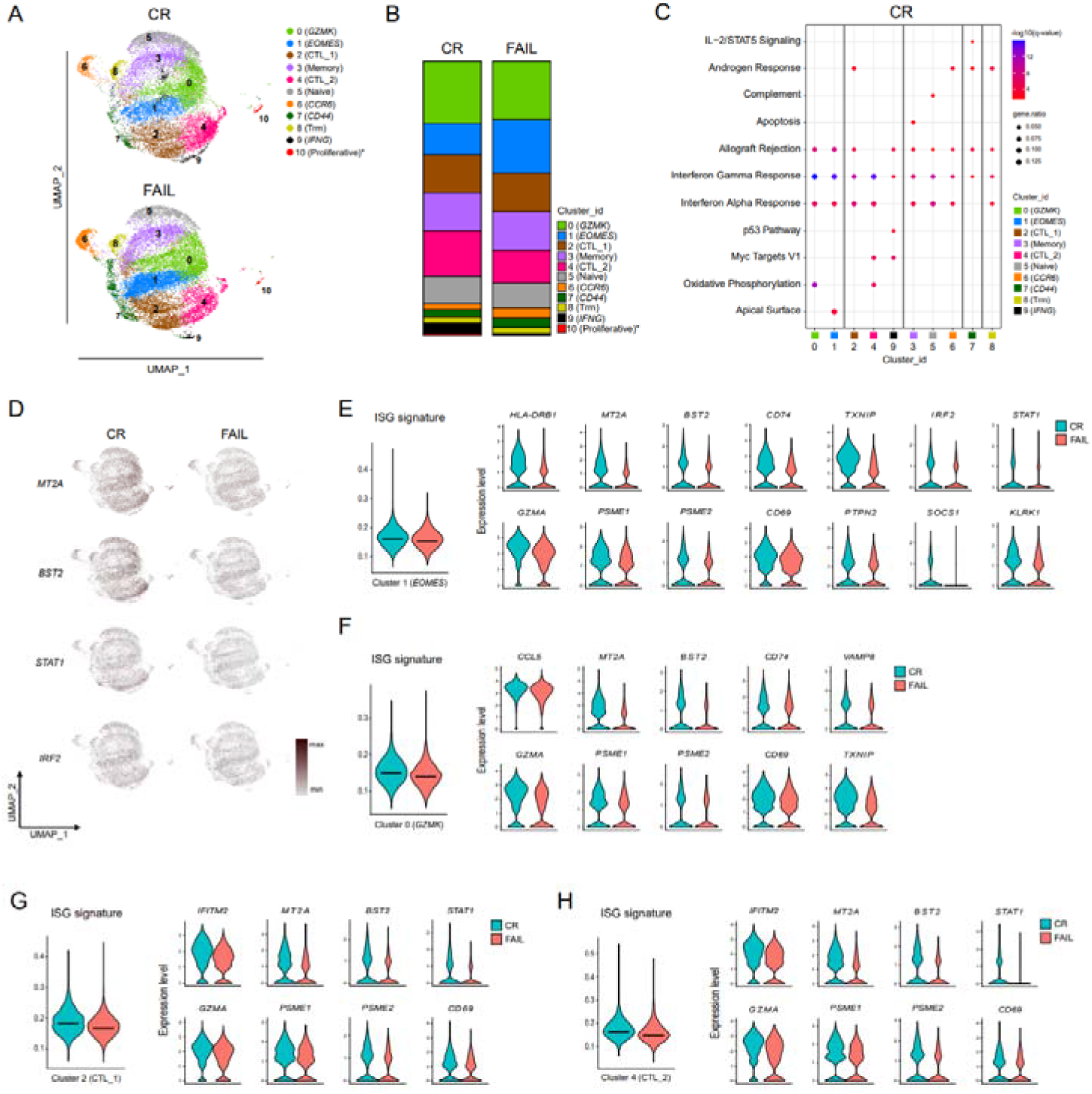
BM-derived CD8^+^ T cells from responders (CR) to AZA show an enhanced ISG molecular signature compared to non-responders (FAIL) in scRNAseq analysis. (**A**) Comparison of separate UMAPs for CR (a total of 13,667 cells, 7,571 from MDS and 6,096 cells from secondary AML patients, respectively) and FAIL patients (a total of 14,782 cells, 8,026 cells from MDS and 6,756 cells from secondary AML patients, respectively). (**B**) Stacked bar chart showing the average distribution of clusters between the two groups. The percentage of cluster 10 (Proliferative) was increased in CR compared to FAIL patients (Unpaired Student’s t test, p=0.0418). (**C**) Dot plot representing MSigDB (Hallmark 2020) enrichment analysis of positively enriched pathways in CR patients. Enriched pathways with a q-value <0.05 (Benjamini-Hochberg correction) are shown. (**D**) Gradient expression of representative selected genes involved in IFN-related pathways, as they are projected onto UMAPs. (**E**) ISG (Interferon Stimulated Genes) score of cluster 0 (*GZMK*) and violin plots showing the expression levels of the top differentially expressed IFN-stimulated genes of cluster 0 (*GZMK*) between CR and FAIL. (**F**) ISG score of cluster 1 (*EOMES*) and violin plots showing the expression levels of the top differentially expressed IFN-stimulated genes of cluster 1 (*EOMES*) between CR and FAIL. (**G**) ISG score of cluster 2 (CTL_1) and violin plots displaying the expression levels of the top differentially expressed IFN-stimulated genes of cluster 2 (CTL_1) between CR and FAIL. (**H**) ISG score of cluster 4 (CTL_2) and violin plots displaying the expression levels of the top differentially expressed IFN-related genes of cluster 4 (CTL_2) between CR and FAIL. Differential expression data are reported in Data sheet 3.

Pathway analysis was performed, and DEGs upregulated in the CR group were associated with IFN response in all clusters (Fig. 6C). To this direction, increased expression of the IFN-responsive genes *MT2A*, *BST2*, *STAT1* and *IRF2* was observed in the CR group (Fig. 6D). We then focused on specific clusters, and we used a score that derives from the expression of interferon stimulated genes (ISG signature).^32^ We observed higher ISG signature score in the CR group in cluster 0 (*GZMK*), which was associated with increased expression of the IFN-responsive genes *CCL5*, *MT2A*, *BST2*, *CD74*, *GZMA*, *PSME1*, *PSME2*, *CD69*, *VAMP8* and *TXNIP* (Fig. 6E). Similarly, increased expression of *HLA-DRB1*, *MT2A*, *BST2*, *CD74*, *TXNIP*, *GZMA*, *PSME1*, *PSME2*, *CD69*, *PTPN2*, *IRF2*, *STAT1*, *SOCS1* and *KLRK1* was observed in cluster 1 (*EOMES*) (Fig. 6F). Regarding the cytotoxic clusters 2 (CTL_1) (Fig. 6G, Supplementary Fig. 12A) and 4 (CTL_2) (Fig. 6H, Supplementary Fig. 12B), higher ISG signature and increased expression of the genes *IFITM2*, *MT2A*, *BST2*, *STAT1*, *GZMA*, *PSME1*, *PSME2* and *CD69* was detected in cells from the CR group from both clusters.

Pathway analysis of DEGs upregulated in the FAIL group demonstrated that cells from the clusters 0 (*GZMK*), 1 (*EOMES*), 2 (CTL_1), 3 (Memory), 4 (CTL_2), 5 (Naïve), 6 (*CCR6*) and 8 (Trm) were enriched for DEGs associated with TNF signaling (Fig. 7A, Supplementary Fig. 13). Moreover, cells from the cytotoxic clusters 2 (CTL_1), 4 (CTL_2), 9 (*IFNG*) and clusters 3 (Memory), 6 (*CCR6*) and 7 (*CD44*) were enriched for DEGs of the TGF-β signaling pathway (Fig. 7A). Specifically for cluster 2 (CTL_1), there was higher TGF-β signature score in the FAIL group and increased expression of *TFGB1*, *SMURF2*, *SMAD7*, *SKI*, *SKIL* (Fig. 7B, Supplementary Fig. 12A), whereas for cluster 4 (CTL_2) the higher TGF-β signature score was associated with increased expression of *TFGB1*, *ARID4B*, *SMAD7*, *SKI*, *SKIL* and *SMURF2* (Fig. 7C, Supplementary Fig. 12B). Since TGF-β signaling has been previously associated with decreased cytotoxicity,^33^ we further used the cytotoxic signature score (Fig. 7D) to address whether the TGF-β signature is associated with decreased cytotoxic activity in CTL clusters in the FAIL group. We observed higher cytotoxicity score in the CR group in cluster 2 (CTL_1), which was associated with the enhanced expression of *GZMA*, *GZMB*, *CX3CR1* and *CCND3* (Fig. 7E) and in cluster 4 (CTL_2), which was associated with enhanced expression of *FGFBP2*, *GZMA*, *GZMB*, *GZMH*, *CCND3*, *C1orf162* and *CX3CR1* (Fig. 7F).

**Fig. 7.**
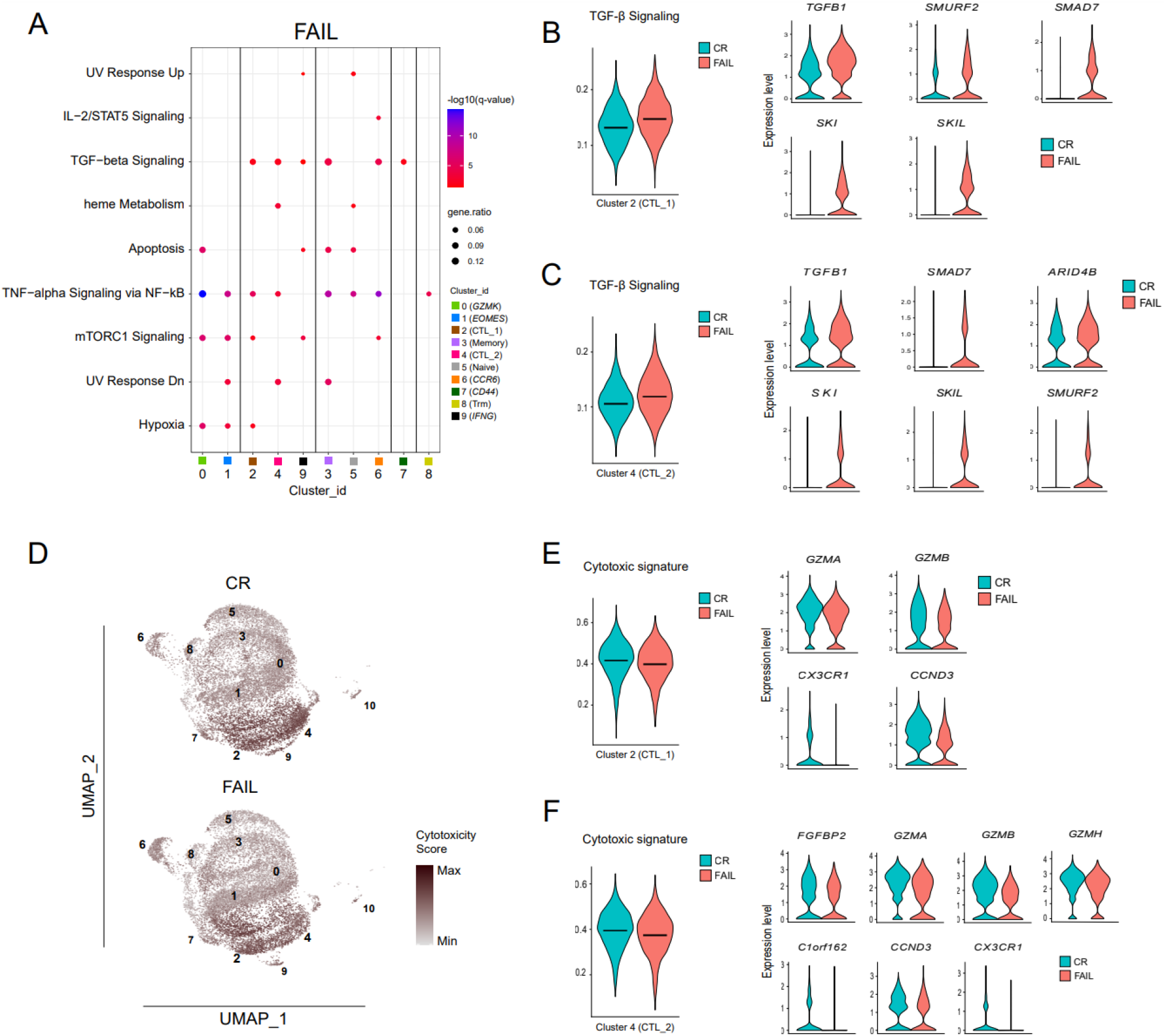
BM-derived CD8^+^ T cells of non-responders (FAIL) displayed suppressed cytotoxic molecular signature at the single-cell level. (**A**) Dot plot representing MSigDB (Hallmark 2020) enrichment analysis of positively enriched pathways in FAIL patients. Enriched pathways with a q-value <0.05 (Benjamini-Hochberg correction) are shown. (**B**) TGF-β signaling score of cluster 2 (CTL_1) and violin plots displaying the expression levels of the top differentially expressed genes of cluster 2 (CTL_1), involved in the enrichment of the TGF-β signaling pathway between CR and FAIL. (**C**) TGF-β signaling score of cluster 4 (CTL_2) and violin plots displaying the expression levels of the top differentially expressed genes of cluster 4 (CTL_2), involved in the enrichment of the TGF-β signaling pathway between CR and FAIL. (**D**) Comparison of the cytotoxic score of each group, as it is projected onto the respective UMAPs. (**E**) Cytotoxic score of cluster 2 (CTL_1) and violin plots exhibiting the expression levels of the top differentially expressed cytotoxicity-related genes of cluster 2 (CTL_1) between CR and FAIL. (**F**) Cytotoxic score of cluster 4 (CTL_2) and violin plots exhibiting the expression levels of the top differentially expressed cytotoxicity-related genes of cluster 4 (CTL_2) between CR and FAIL. Differential expression data are reported in Data sheet 3.

We further utilized the dysfunctional score^31^ to study potential differences in the expression of exhaustion-associated genes based on treatment outcome. This analysis revealed no significant differences in the dysfunctional score across all clusters between CR and FAIL patients (Supplementary Fig. 14).

Cytokines act together with transcription factors (TFs) to regulate T cell fate and functionality.^34^ To study the TFs that could act as possible regulators of the transcriptomic alterations observed in responders to AZA compared to non-responders, TF regulatory network analysis was performed using SCENIC,^35^ which resulted in the identification of 11 clusters-regulons (Fig. 8A). We observed that regulon 5 was enriched with cells from the CR group of patients, whereas regulon 7 was enriched with cells from the FAIL group (Fig. 8B, C). Regulon cluster 5 was characterized by IRF7-, CHURC1-, NFYB- and RFXANK-regulated networks (Fig. 8D) and was enriched mainly with cells from cluster 4 (CTL_2) (Supplementary Fig. 15). On the other hand, regulon 7 was characterized by the NFKB2-, REL-, RELB-, CREM- and NFKB1-regulated networks (Fig. 8D) and it was enriched with cells from cluster 0 (*GZMK*) and cluster 1 (*EOMES*) (Supplementary Fig. 15), previously described as pre-dysfunctional cells.^25,26^

**Fig. 8.**
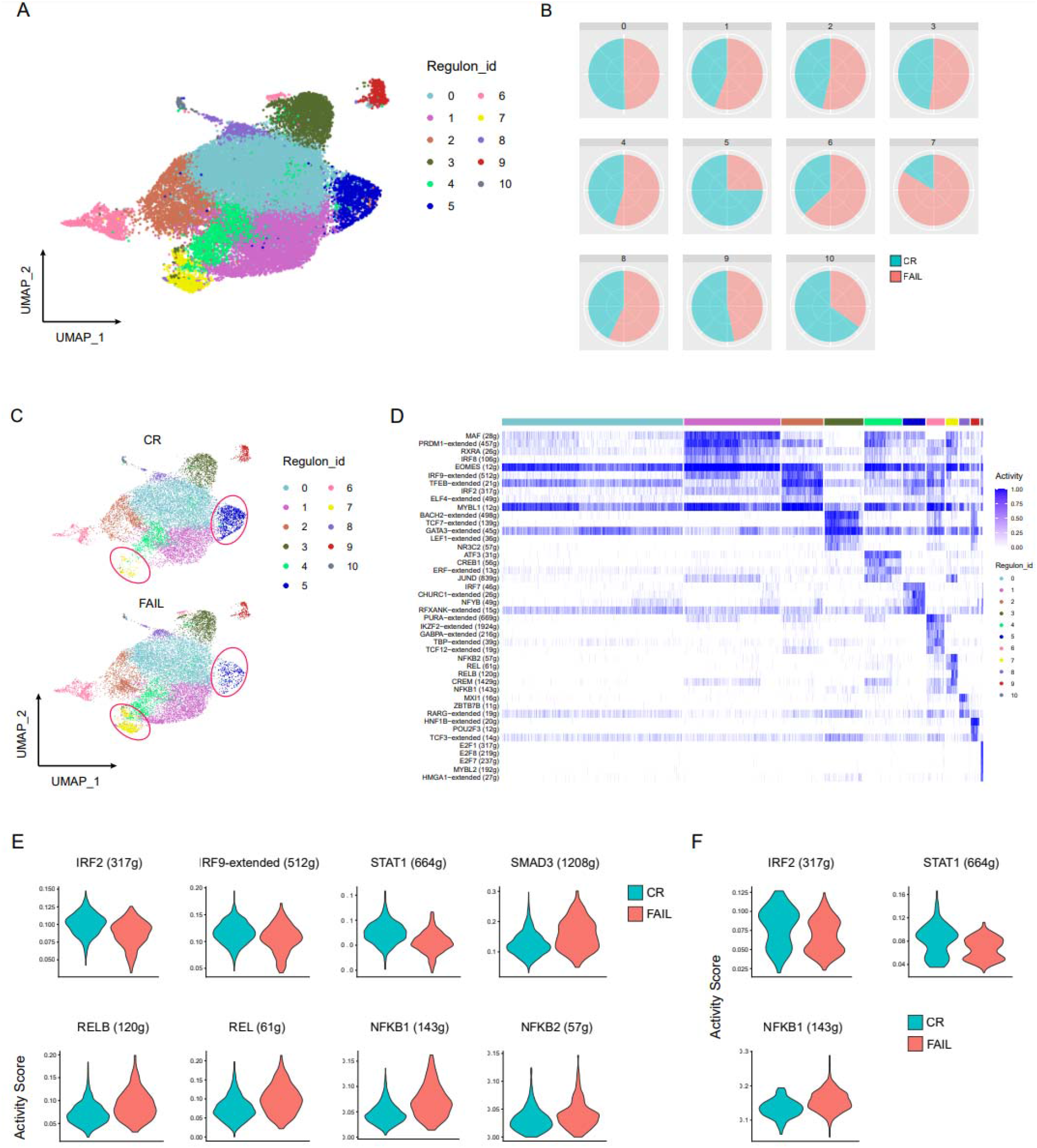
Transcription Factor (TF) regulatory network analysis in BM-derived CD8^+^ T cells. (**A**) UMAP depicting the clustering of CD8^+^ T cells based on regulons. (**B**) Pie charts illustrating the representation of cells from CR and FAIL patients within each regulon. (**C**) Comparison of cell distribution in regulons between the groups utilizing separate UMAPS for each group. (**D**) Heatmap showing the top differentially activated TFs of each regulon-cluster. (**E-F**) Violin plots depicting the activity score of selected TFs per sample type in regulons 5 and 7, respectively.

Differential TF activity analysis between the two groups of patients within regulon 5, predicted that the activity of the interferon-related TFs IRF2, IRF9 and STAT1 was increased in the CR group and the activity of SMAD3, and the NF-κB superfamily TFs RELB, REL, NFKB2 and NFKB1 was increased in the FAIL group (Fig. 8E). Similar analysis within regulon 7, predicted increased activity for IRF2 and STAT1 and decreased activity for NFKB1 in cells derived from the CR group (Fig. 8F). Taken together, the TF regulatory network analysis further supports that BM CD8^+^ T cells in the CR group are targeted by TFs associated with IFN signaling, whereas TFs linked to TGF-β and NF-κB signaling target CD8^+^ T cells in the FAIL group.

## Discussion

Alterations in CD8^+^ T cell functionality in the tumor milieu promote tumor evasion and compromise response to immunotherapies.^25^ In AML and MDS, several CD8^+^ T cell defects have been described that are potentially reversible by various treatments, including AZA.^9,11,36^ However, the architecture of CD8^+^ T cell immunity and its impact on the clinical behavior of AML and MDS are still incompletely understood, thus hampering the development of successful immunotherapeutic approaches.

Herein, we provide a systemic analysis at the single cell level of CD8^+^ T cells derived from the BM immune microenvironment of patients with clonal myeloid disorders. We specifically focused on the identification of specific immunophenotypic, and molecular signatures associated with the outcome of AZA treatment.

Deep immunophenotyping with CyTOF resulted in the identification of a BM CD57^+^CXCR3^+^CD8^+^ T cell population with increased frequency in patients with AML and CMML compared to MDS. We further observed that increased pre-treatment frequency of BM CD57^+^CXCR3^+^CD8^+^ T cells was associated with poor OS and response to AZA treatment in patients with HR-MDS and AML. Interestingly, no association between the frequency of this cell population and disease course was observed in patients with CMML, supporting the notion that this disorder does not share common immune features with MDS and AML. Though the mutational profile may sculpt specific immune response patterns across heterogeneous tumors,^37^ no definite association between somatic mutations and defects in T cell immunity has been shown in MDS.^38^ Consistent with this report and our previous observation, showing absence of an association between alterations in CD4^+^ cells and somatic mutations,^21^ we could not find any correlation between the levels of the aforementioned subpopulation with the mutational profile of patients.

The expression of CD57 by CD8^+^ T cells, coupled with the absence of CD28 expression, characterizes a senescent-like phenotype, linked to chronic immune activation in several disorders.^12,39,40^ These CD57^+^CD28^-^CD8^+^ T cells are antigen specific effector cells that have limited proliferation capacity, due to their advanced differentiation stage.^41^ Previous studies demonstrated increased levels of CD57^+^CD28^-^CD8^+^ T cells in patients with MDS and AML compared to healthy individuals.^10,42^ CXCR3, on the other hand, is expressed on Th1-CD4^+^ T cells^43^ and effector CD8^+^ T cells and is considered crucial for the recruitment of T cells to inflammatory sites.^44^ The expression of CXCR3 on CD8^+^ T cells has been associated with changes in the equilibrium from memory towards effector cell populations.^45^ Interestingly, recent studies engaging scRNA-seq in CD8^+^ T cells isolated from patients with cancer have described cells expressing *CXCR3* together with *GZMK* and *EOMES* as a pre-dysfunctional cell population.^26^

In line with our findings, it has been recently demonstrated that the accumulation of senescent-like CD8^+^CD57^+^ T cells is negatively associated with the response to chemotherapy and checkpoint blockade immunotherapy in AML patients.^46^ By utilizing *in vitro* studies, the authors further demonstrated that patient-derived senescent-like T cells were not able to sufficiently eliminate autologous AML-blasts when compared to their non-senescent CD8^+^ T cell counterparts, providing direct evidence for their limited antileukemic activity.^46^

Based on our findings from immunophenotyping, we engaged scRNA-seq analysis to investigate in depth the transcriptomic profile of BM CD8^+^ T cells from patients with HR-MDS and secondary AML. We observed that the abundance of cells within the CTL_2 cluster, characterized by higher expression compared to other clusters of *B3GAT1*, the gene that encodes the CD57 protein, was increased in patients with secondary AML compared to HR-MDS, being in line with the immunophenotypic findings. Regarding response to treatment, a significant enrichment of the TGF-β signaling pathway was observed in cytotoxic clusters (CTL_1 and CTL_2) of non-responders. This pathway has been previously shown to directly inhibit the cytotoxic program of CD8^+^ T cells leading to compromised anti-tumor responses, tumor evasion and poor outcomes.^33^ In line with this, non-responders displayed decreased cytotoxic signature within the same clusters. Of note, upregulated TGF-β signaling has also been reported in *ex vivo*–expanded BM mesenchymal cells from AZA-treated patients,^47^ potentially implying a ubiquitous targeting of TGF-β signaling by AZA. Luspatercept, an inhibitor of TGF-β signaling, has been recently approved by the FDA for the treatment of transfusion dependent LR-MDS patients either after failure to erythropoiesis stimulating agents or at first line.^48^ The effect of luspatercept on tumor immunity is still unknown, but, given the immunoregulatory role of TGF-β, it could be worthwhile considering the potential benefit of adding luspatercept in AZA refractory patients.

On the other hand, responders exhibited a significant enrichment of pathways associated with IFN response, across many BM CD8^+^ T cell clusters. Interferons play a crucial role in immune activation during anti-tumor responses.^49,50^ Notably, there is evidence indicating that AZA exerts its effects in an IFN-dependent manner by increasing the production of IFN-γ in T cells,^51^ and by activating type I and III Interferon signaling in tumor cell lines,^52,53^ as well as upregulating the expression of Interferon Stimulated Genes (ISGs), all of which aid in the rejuvenation of the anti-tumor immunity.^52^ Taken together, our findings suggest that the balance between TGF-β and IFN signaling in CTLs within the BM microenvironment may regulate their anti-tumor effect, affecting the response to treatment.

Immunotherapies based on immune checkpoint blockade (ICB) have revolutionized the field of oncology.^54^ Many studies employing scRNA-seq in solid tumors demonstrated that CD8^+^ T cells show a distinct molecular signature, indicative of an exhausted phenotype, which is associated with disease progression and treatment resistance,^31,55^ thereby providing evidence supporting the effectiveness of ICB treatment in these patients. In contrast, ICB therapy in myeloid malignancies, including MDS and AML, has yielded limited results so far, while the risk of serious adverse events still remains.^56^ For this reason, the identification of alternative pathways, such as TGF-β signaling pathway, that could be targeted by immunotherapies in patients with myeloid neoplasms is of paramount importance.

In conclusion, by performing a mass cytometry-guided transcriptomic analysis of BM CD8^+^ T cells at the single cell level we provide evidence of predictive abnormalities of BM CD8^+^ cells in AML and MDS patients treated with AZA. We further identified TGF-β signaling in BM CD8^+^ T cells as a potential immune-mediated mechanism of resistance to AZA, thus arguing for the use of inhibitors of the TGF-β pathway to prevent or overcome AZA refractoriness. In view of recent data suggesting a T-cell mediated antileukemic activity of venetoclax,^36^ a triple combination of AZA, venetoclax and luspatercept may have the potential to induce a fully competent immune-mediated control of the leukemic clone in AML and MDS patients.

## Methods

### Study design

The overall aim of our study was to investigate the BM immune landscape of patients with myeloid neoplasms, aiming to uncover specific immunophenotypic disparities and molecular signatures related to the development and advancement of myeloid neoplasms, as well as the response of patients to AZA treatment. To this direction, we initially engaged mass cytometry (CyTOF) to study the immune cell populations of BM samples from patients with LR-MDS, HR-MDS, CMML and AML collected before treatment initiation. We, then, engaged an additional cohort and performed flow cytometry, to validate the results derived from CyTOF. Based on the findings from the immunophenotypic analysis we focused on BM CD8^+^ T cells and performed scRNA-seq, to address the molecular signature in CD8^+^ T cell subsets.

### Study patients

BM samples were collected from treatment-naïve patients with MDS, AML and CMML-2. Patient diagnosis was conducted based on the 2022 5th World Health Organization (WHO) classification^57^ and MDS patients were categorized based on their IPSS-R score, into lower-risk MDS (LR-MDS; IPSS-R score ≤ 3.5) and higher-risk MDS (HR-MDS; IPSS-R score > 3.5).^58^ Except from patients with LR-MDS, patients were treated with Azacitidine (AZA) in a subcutaneous dose of 75 mg/m^2^ for 7 consecutive days within a 28-day cycle. To manage myelotoxicity or complications associated with myelosuppression, potential measures such as reducing the dose by up to 50% or delaying treatment were considered. Response assessment was determined using the International Working Group Response Criteria for MDS^59^ and the recently revised European LeukemiaNet criteria for AML.^60^ Detailed demographic and clinical data are presented in Table 1. The study was approved by the local Ethics Committee, under the reference number (877/23-10-2019). All patients provided informed written consent in accordance with the principles outlined in the Declaration of Helsinki.

**Table 1.** Baseline patient characteristics.

|  |  |
| --- | --- |
| **Total No of patients, N** | 104 |
| **Age at diagnosis, N (%)** |  |
| >65yo | 77 (74%) |
| <65yo | 22 (21,2%) |
| N/A | 5 (4,8%) |
| **Sex, N (%)** |  |
| Male | 69 (66.35%) |
| Female | 35 (33.65%) |
| **WHO classification, N (%)** |  |
| MDS-LB | 19 (18.3%) |
| MDS-IB1 | 15 (14.4%) |
| MDS-IB2 | 26 (25.0%) |
| AML | 32 (30.8%) |
| CMML-2 | 12 (11.5%) |
| **Baseline blood counts, median (range)** |  |
| Hemoglobin (g/dL) | 9.4 (5.4-14) |
| Absolute neutrophil count; ANC ($\times 10^9/L$) | 2.01 (0-60) |
| Platelet count ($\times 10^9/L$) | 90.5 (5-463) |
| **(%) Bone marrow blasts, N (%)** |  |
| <5% | 17 (16,4%) |
| 5-10% | 20 (19.2%) |
| 11-20% | 30 (28,85%) |
| >20% | 30 (28,85%) |
| N/A | 7 (6,7%) |
| **Cytogenetic risk score (IPSS-R), N (%)** |  |
| Very good | 3 (2,9%) |
| Good | 57 (54,8%) |
| Intermediate | 25 (24%) |
| Poor | 2 (1,9%) |
| Very poor | 9 (8,7%) |
| N/A | 8 (7,7%) |
| **Number of completed AZA cycles, median (range)** |  |
| HR-MDS | 9 (1-59) |
| AML | 7(1-70) |
| CMML-II | 7 (3-36) |

### Collection and handling of samples

Density gradient centrifugation, using Ficoll-Histopaque 1077 (Sigma-Aldrich), was employed to isolate bone marrow mononuclear cells (BMMCs). Immediately after isolation, BMMCs were cryopreserved in a freezing medium consisting of 90% Fetal Bovine Serum (FBS) and 10% Dimethyl Sulfoxide (DMSO).

### Mass cytometry and data analysis

High-dimensional immunophenotyping of BMMCs was performed with mass cytometry using established and validated workflows from previous studies.^61,62^ We employed the Maxpar Direct Immune Profiling Assay (MDIPA) that contains 30 pre-conjugated antibodies (Supplementary Table 3) with metal probes in lyophilized form (Standard Biotools).^63^ Prior to staining, BMMCs were thawed in prewarmed RPMI medium and supplemented with 10% FBS. After two washes, the cells were resuspended in fresh medium. BMMCs were subjected to a blocking step using Human TruStain FcX (Biolegend). Subsequently, cells were stained for surface markers following the MDIPA manufacturer’s instructions. Two additional washes with Cell Staining Buffer (CSB) were performed, and fixation was carried out using a 1.6% filtered formaldehyde solution from Sigma for 20 minutes at room temperature. Finally, cells were stained in a DNA intercalator solution (1:1000 dilution of 125 μM Cell-ID™ Intercalator-Ir) in Maxpar Fix and Perm buffer (Standard BioTools). The next day, cells were washed with CSB buffer and Cell Acquisition Solution (CAS) and then resuspended with EQ Passport beads (Standard Biotools, 1:10 dilution) immediately before acquisition. Acquisition was performed using a Helios™ system. To ensure data quality during acquisition, the flow rate at the Helios™ system did not exceed 350 events per second. Data were subsequently normalized using Passport beads with CyTOF software (version 10.7.1014). Prior to analysis, we performed data cleanup, with bivariate dot plots in FlowJo™ (v10.8 Software, BD Biosciences), to refine gaussian parameters, and live, singlet cell events were selected for downstream analysis. Data analysis was performed on CD45^+^ cells, to exclude blasts from the analysis. FlowSOM clustering analysis and dimensionality reduction via tSNE were carried out in the R programming environment (version 4.1.0), following established open-source workflows previously described.^62^

For targeted analysis of T cell populations, data were imported into Cytobank (accessible at https://premium.cytobank.org) for further assessment. All related statistical tests and illustrations were generated through Cytobank. The FlowSOM algorithm was utilized to hierarchically cluster gated CD4^+^ and CD8^+^ T cell populations into distinct metaclusters, based on their surface marker expression profiles. Proportional sampling was employed to maximize the inclusion of total events in the analysis. The default/automatic settings were used for the clustering method, iterations, seed, and number of clusters, while the number of metaclusters was set to 10 or 6 based on the specific analysis requirements. For illustration purposes, metaclusters were also projected onto representative tSNE maps that were generated using the dimensionality reduction algorithm tSNE-CUDA.

### Flow cytometry

Sample preparation and flow cytometry was performed as previously described.^21^ Details about the antibodies used are provided in Supplementary Table 4. Data were collected, on an 8-color flow cytometer FACS Canto II (BD Biosciences), using BD FACSDiva^TM^ (version 8.0.1 for Windows) software and subsequently analyzed using FlowJo^TM^ (version 10 for Windows, BD Biosciences) software.

### Next-generation sequencing

DNA extraction was performed on BMMCs or peripheral blood mononuclear cells (PBMCs), before treatment initiation. The genomic DNA was isolated utilizing the Purelink Genomic DNA Mini Kit (Invitrogen, #K182001). The VariantPlex Myeloid panel was utilised to detect copy number variations (CNVs), single-nucleotide variants (SNVs) and indels in 75 myeloid associated genes as per manufacturer’s instructions. Target-enriched libraries of extracted nucleic acids for next-generation sequencing were prepared using Anchored Multiplex PCR (AMP), a target enrichment chemistry utilizing unidirectional gene-specific primers (GSPs), sample indexes and molecular barcodes for multiplex targeted NGS. After sequencing on an Illumina platform, analysis was performed on Archer Analysis bioinformatics platform.

### scRNA-seq and data processing

BMMCs were thawed and washed with RPMI-1640 (GlutaMAX™, Gibco, #61870) supplemented with 10% heat-inactivated fetal bovine serum (FBS, Gibco, #10270), 100 U/ml penicillin–streptomycin (10,000 U/ml, Gibco, #15140). Then, BMMCs were treated with DNAse I 1mg/ml (0.25 mg ml−1, Sigma) for 10 min at room temperature. Samples were stained with 7-AAD Viability Staining Solution (420404, Biolegend), CD45 APC/Cy7 (304014, Biolegend), CD3 PE (317308, Biolegend), CD8 APC (345775, BD), CD4 FITC (345768, BD Biosciences). After antibody staining, cells were incubated with Cell Multiplexing Oligos (3’ CellPlex Kit Set A, 10x Genomics) following the manufacturer’s instructions. Based on 7-AAD-CD3^+^CD4^-^CD8^+^ profile, cells were sorted on a FACS ARIA III (BD Biosciences) v8.0.1 software (BD Biosciences). Cell purity was above 95%.

Sorted cells were counted, resuspended in PBS + 10% FBS at a concentration of 1600 cells/ uL, combined by three per well of the Chromium Next GEM Chip G (10X Genomics) and loaded onto the Chromium Controller (10X Genomics). Samples were processed for single-cell encapsulation, cDNA and cell multiplexing library generation using the Chromium Next GEM Single Cell 3LJ Reagent Kits v3.1 (Dual Index) (10X Genomics). The constructed libraries were sequenced on an NovaSeq 6000 sequencer with a paired-end reads sequencing mode. The 10X Genomics Cell Ranger multi v7.1.0 pipeline, was used to map the sequencing reads to the human genome (GRCh38) and generate the gene expression and feature barcode matrices. We specified the r1-length and r2-length to 28+90, respectively, for both the gene expression and feature libraries, while preserving all others parameters of the pipeline under default setting. The generated matrices consisted of 38298 cellular barcodes, spanning all samples, with a sequencing depth of 34822 mean reads per barcode (cDNA) and were inserted to the R (Version 4.1.1) software package Seurat (v4.3.0)^64^ for all downstream analyses. The gene expression matrices were filtered to discard cells expressing less than 200 genes as well as genes found in less than 3 cells. Each library was processed individually for sample demultiplexing and singlet identification in R using Seurat “HTODemux” function based on the feature barcode matrices generated by CellRanger. Finally, cellular barcodes corresponding to single cells were merged to a single Seurat object. During quality control of the combined dataset, cells expressing less than 200 or more than 4500 genes and having more than 13% of mitochondrial associated genes were removed from further analysis. Gene expression data of the remaining 28585 cells that passed quality control, were normalized and scaled using the “LogNormalize” method and “ScaleData” command, respectively, while variable features were identified using the “FindVariableFeatures” command. The algorithm Harmony^65^ was used to perform batch correction and for further clustering of the data. The first 63 principal components of the Harmony reduction were selected, based on the Seurat Elbow plot, and were designated for the “dims” argument of the “FindNeighbors” and “RunUMAP” functions. A resolution of 0.3 was selected for graph-based cluster identification in the “FindClusters” function and, finally, cluster visualization in a two-dimensional space was performed using non-linear dimensional reduction via uniform manifold approximation and projection (UMAP). The “FindAllMarkers” command was implemented to identify cluster defining genes, expressed at least in 20% of the cluster cells at a minimum of 0.25-log-fold difference between the respective cluster and the residual cells in the dataset. Contaminating clusters, comprising non-CD8^+^ T cells were identified via their gene expression profile and were removed from subsequent analysis. To identify differentially expressed markers between conditions in the same cluster(s) the “FindMarkers” command with the MAST statistical test were used. For increased sensitivity purposes, the output of the “FindMarkers” comparison contained genes having an adjusted p-value < 0.05, expressed in at least 5% of the cells in least one condition, without implementing log-fold-change thresholds. Calculation of curated gene set scores was performed with the AUCell package.^35^

The identification of genes displaying statistically significant differential expression was conducted by applying an adjusted p value (q-value) threshold of <0.05 and a |log2 fold change (logFC)| threshold of >0.25. Subsequently, an enrichment pathway analysis of the differentially expressed genes was performed using the EnrichR tool.^66,67^ Trajectories were performed using Monocle 3 using default settings.^68^

### Gene regulatory network analysis

The single-cell RNA-seq data underwent further bioinformatic analysis through the utilization of SCENIC. SCENIC is a computational tool that constructs intricate gene regulatory networks (GRNs) and uncovers distinct cellular states within the framework of single-cell RNA-seq data.^35^ SCENIC relies on three main R/bioconductor packages: GENIE3, RcisTarget, and AUCell. GENIE3^69^ identifies potential targets of Transcription Factors (TFs) by elucidating the coexpression relationships existing between these TFs and their corresponding targets. Subsequently, RcisTarget^35^ is utilized to pinpoint direct targets through the analysis of cis-regulatory motifs and construct transcription factor regulons. The third component, AUCell,^35^ is used to assess the activity of each of these regulons within individual cells. SCENIC was employed to establish regulons, estimate the activity scores of transcription factors, as well as to conduct enrichment analysis. Additionally, differential regulon activity analysis was conducted within the clusters previously identified by Seurat. The outcomes were represented through UMAPs, heatmaps and pie charts. Notably, the heatmap that depicts the activity of certain regulons per cluster (switched on/off state) was based on the binary values generated by the AUCell algorithm. The methodology and the scripts for this part used as a guide the SCENIC vignettes curated by Aibar et al,^35^ along with relative scripts by Zhu et al.^70^

### Statistical analysis

Τo compare two groups, a two-tailed unpaired Student’s t-test or a Mann-Whitney U test was used as appropriate. For the comparison of multiple groups, one-way ANOVA followed by a “two-stage” Benjamini, Krieger & Yekutieli multiple comparison test, or Kruskal-Wallis test followed by a “two-stage” Benjamini, Krieger & Yekutieli multiple comparison test were used as appropriate. The independence between variables was assessed using the chi-square (χ²) test and Fisher’s exact test. Kaplan-Meier analysis was utilized for survival analysis, and the log-rank test was employed to assess the significance. To explore the association of BM CD57^+^CXCR3^+^CD8^+^ T cells with response to treatment, univariate analysis was performed. Optimal CD57^+^CXCR3^+^ cut-off was determined by transformation of scale variable to binary one through optimal scaling; in detail, discretization to seven groups, regularization using ridge regression, and 10-fold cross-validation were performed through SPSS CATREG procedure. To explore the independent correlations between BM CD57^+^CXCR3^+^CD8^+^ T cells along with mutational status, as well as age, gender, and IPSS-R (both score and components), with response to treatment, a single univariate and separate multivariate analyses for each mutation of interest were performed using Cox proportional hazards regression analysis; every parameter that was significantly correlated in a certain univariate analysis (p≤0.05) was treated as a potential independent parameter in the relevant multivariate one. Statistical analysis was performed using GraphPad prism 9 (version 9.0.0 for Windows, GraphPad Software, Inc., San Diego, CA, USA.), IBM SPSS Statistics software (version 26.0 for Windows, IBM Corporation, North Castle, NY, USA) and the Cytobank platform. The level of significance was established at P < 0.05.

## Data availability

The authors state that all data supporting this study are available in the main text or the supplementary material. The raw scRNA-sequencing data for this study have been deposited in the NCBI Gene Expression Omnibus repository and are accessible through accession number GSE250077. All other raw data can be provided by the authors upon reasonable request.

## Funding

This study was supported by the Hellenic Foundation for Research and Innovation (HFRI) under the HFRI ‘’Research Projects to Support Faculty Members & Researchers and Procure High-Value Research Equipment’’ grant (HFRI-FM17-452) and the General Secretariat for Research and Technology Management and Implementation Authority for Research, Technological Development, and Innovation Actions (MIA-RTDI) (grant T2EDK-02288, MDS-TARGET). IM was funded by the RIG-2023-051 grant from Khalifa University. TA and NEP are supported by the ERC under the European Union’s Horizon 2020 research and innovation program (grant agreement no 947975 to T. Alissafi) and from the Hellenic Foundation for Research and Innovation (H.F.R.I.) under the “2nd Call for H.F.R.I. Research Projects to support Post-Doctoral Researchers” (Project Number: 166 to T. Alissafi) and the “Sub-action 1. Funding New Researchers – RRF: Basic Research Financing (Horizontal support for all Sciences) (Project number: 15014 to T. Alissafi). IPK is funded by the Academy of Medical Sciences (R2429101) and Rosetrees Trust (R2449101). TC is supported by the Deutsche Forschungsgemeinschaft (TRR332, project B4).

## Acknowledgments

AT presented this study at the 34^th^ Meeting of Hellenic Society of Haematology and was awarded with the “A. Goutas” award.

## Authors’ contributions

Conceptualization: AT, IK, IM

Methodology: AT, MG, NEP, NP, PC, TC, TA, IM

Investigation: AT, MG, KLO, AF, KK, EL, VP, AC, IPK, PG, PK, PL, IKY, KL

Visualization: AT, NEP, NP

Funding acquisition: IPK, TC, TA, IM

Project administration: IM

Supervision: IM

Writing – original draft: AT, IM

Writing – review & editing: AT, IPK, TC, IK, IM

## Competing interests

The authors declare no competing financial interests.

## Notes

### Competing Interest Statement

The authors have declared no competing interest.

### Author Declarations

The study was approved by the Ethics Committee of the University Hospital of Alexandroupolis, under the reference number (877/23-10-2019).

### Summary of Updates

Addion of figure 5, describing a further analysis of the data.

## References

1. Arber, D. A. et al. International Consensus Classification of Myeloid Neoplasms and Acute Leukemias: integrating morphologic, clinical, and genomic data. Blood 140, 1200–1228 (2022).

2. Ogawa, S. Genetics of MDS. Blood 133, 1049–1059 (2019).

3. Itzykson, R. & Fenaux, P. Epigenetics of myelodysplastic syndromes. Leukemia 28, 497–506 (2014).

4. Aggarwal, S. et al. Role of immune responses in the pathogenesis of low-risk MDS and high-risk MDS: implications for immunotherapy. Br J Haematol 153, 568–581 (2011).

5. Balderman, S. R. et al. Targeting of the bone marrow microenvironment improves outcome in a murine model of myelodysplastic syndrome. Blood 127, 616–625 (2016).

6. Kitagawa, M. et al. Localization of Fas and Fas ligand in bone marrow cells demonstrating myelodysplasia. Leukemia 12, 486–492 (1998).

7. Pronk, E. & Raaijmakers, M. H. G. P. The mesenchymal niche in MDS. Blood 133, 1031–1038 (2019).

8. Verma, N. K. et al. Obstacles for T-lymphocytes in the tumour microenvironment: Therapeutic challenges, advances and opportunities beyond immune checkpoint. EBioMedicine 83, 104216 (2022).

9. Knaus, H. A. et al. Signatures of CD8+ T cell dysfunction in AML patients and their reversibility with response to chemotherapy. JCI Insight 3, 120974 (2018).

10. Radpour, R. et al. CD8+ T cells expand stem and progenitor cells in favorable but not adverse risk acute myeloid leukemia. Leukemia 33, 2379–2392 (2019).

11. Rodriguez-Sevilla, J. J. & Colla, S. T cell dysfunctions in myelodysplastic syndromes. Blood blood.2023023166 (2024) doi:10.1182/blood.2023023166.

12. Liu, X., Hoft, D. F. & Peng, G. Senescent T cells within suppressive tumor microenvironments: emerging target for tumor immunotherapy. J Clin Invest 130, 1073–1083.

13. Raskov, H., Orhan, A., Christensen, J. P. & Gögenur, I. Cytotoxic CD8+ T cells in cancer and cancer immunotherapy. Br J Cancer 124, 359–367 (2021).

14. Costa, R. et al. Activity of azacitidine in chronic myelomonocytic leukemia. Cancer 117, 2690–2696 (2011).

15. Platzbecker, U. & Fenaux, P. Recent frustration and innovation in myelodysplastic syndrome. Haematologica 101, 891–893 (2016).

16. Stomper, J., Rotondo, J. C., Greve, G. & Lübbert, M. Hypomethylating agents (HMA) for the treatment of acute myeloid leukemia and myelodysplastic syndromes: mechanisms of resistance and novel HMA-based therapies. Leukemia 35, 1873–1889 (2021).

17. DiNardo, C. D. et al. Azacitidine and Venetoclax in Previously Untreated Acute Myeloid Leukemia. New England Journal of Medicine 383, 617–629 (2020).

18. Prébet, T. et al. Outcome of high-risk myelodysplastic syndrome after azacitidine treatment failure. J Clin Oncol 29, 3322–3327 (2011).

19. Garcia-Manero, G. & Fenaux, P. Hypomethylating agents and other novel strategies in myelodysplastic syndromes. J Clin Oncol 29, 516–523 (2011).

20. Zhao, G., Wang, Q., Li, S. & Wang, X. Resistance to Hypomethylating Agents in Myelodysplastic Syndrome and Acute Myeloid Leukemia From Clinical Data and Molecular Mechanism. Front Oncol 11, 706030 (2021).

21. Lamprianidou, E. et al. Modulation of IL-6/STAT3 signaling axis in CD4+FOXP3− T cells represents a potential antitumor mechanism of azacitidine. Blood Adv 5, 129–142 (2021).

22. Costantini, B. et al. The effects of 5-azacytidine on the function and number of regulatory T cells and T-effectors in myelodysplastic syndrome. Haematologica 98, 1196–1205 (2013).

23. Kornblau, S. M. et al. Recurrent expression signatures of cytokines and chemokines are present and are independently prognostic in acute myelogenous leukemia and myelodysplasia. Blood 116, 4251–4261 (2010).

24. Sand, K. E., Rye, K. P., Mannsåker, B., Bruserud, O. & Kittang, A. O. Expression patterns of chemokine receptors on circulating T cells from myelodysplastic syndrome patients. Oncoimmunology 2, e23138 (2013).

25. van der Leun, A. M., Thommen, D. S. & Schumacher, T. N. CD8+ T cell states in human cancer: insights from single-cell analysis. Nat Rev Cancer 20, 218–232 (2020).

26. Zhang, L. et al. Lineage tracking reveals dynamic relationships of T cells in colorectal cancer. Nature 564, 268–272 (2018).

27. Guo, X. et al. Global characterization of T cells in non-small-cell lung cancer by single-cell sequencing. Nat Med 24, 978–985 (2018).

28. Clarke, J. et al. Single-cell transcriptomic analysis of tissue-resident memory T cells in human lung cancer. J Exp Med 216, 2128–2149 (2019).

29. Szabo, P. A., Miron, M. & Farber, D. L. Location, location, location: Tissue resident memory T cells in mice and humans. Sci Immunol 4, eaas9673 (2019).

30. Kok, L., Masopust, D. & Schumacher, T. N. The precursors of CD8+ tissue resident memory T cells: from lymphoid organs to infected tissues. Nat Rev Immunol 22, 283–293 (2022).

31. Li, H. et al. Dysfunctional CD8 T Cells Form a Proliferative, Dynamically Regulated Compartment within Human Melanoma. Cell 176, 775–789.e18 (2019).

32. Chiche, L. et al. Modular transcriptional repertoire analyses of adults with systemic lupus erythematosus reveal distinct type I and type II interferon signatures. Arthritis Rheumatol 66, 1583–1595 (2014).

33. Batlle, E. & Massagué, J. Transforming Growth Factor-β Signaling in Immunity and Cancer. Immunity 50, 924–940 (2019).

34. Kaech, S. M. & Cui, W. Transcriptional control of effector and memory CD8+ T cell differentiation. Nat Rev Immunol 12, 749–761 (2012).

35. Aibar, S. et al. SCENIC: single-cell regulatory network inference and clustering. Nat Methods 14, 1083–1086 (2017).

36. Lee, J. B. et al. Venetoclax enhances T cell-mediated antileukemic activity by increasing ROS production. Blood 138, 234–245 (2021).

37. Thorsson, V. et al. The Immune Landscape of Cancer. Immunity 48, 812–830.e14 (2018).

38. Winter, S., Shoaie, S., Kordasti, S. & Platzbecker, U. Integrating the ‘Immunome’ in the Stratification of Myelodysplastic Syndromes and Future Clinical Trial Design. J Clin Oncol 38, 1723–1735 (2020).

39. Tae Yu, H., et al. Characterization of CD8(+)CD57(+) T cells in patients with acute myocardial infarction. Cell Mol Immunol 12, 466–473 (2015).

40. Strioga, M., Pasukoniene, V. & Characiejus, D. CD8+ CD28- and CD8+ CD57+ T cells and their role in health and disease. Immunology 134, 17–32 (2011).

41. Brenchley, J. M. et al. Expression of CD57 defines replicative senescence and antigen-induced apoptotic death of CD8+ T cells. Blood 101, 2711–2720 (2003).

42. Epling-Burnette, P. K. et al. Prevalence and clinical association of clonal T-cell expansions in Myelodysplastic Syndrome. Leukemia 21, 659–667 (2007).

43. Yoon, S. H. et al. Selective addition of CXCR3+CCR4-CD4+ Th1 cells enhances generation of cytotoxic T cells by dendritic cells in vitro. Exp Mol Med 41, 161–170 (2009).

44. Maurice, N. J., McElrath, M. J., Andersen-Nissen, E., Frahm, N. & Prlic, M. CXCR3 enables recruitment and site-specific bystander activation of memory CD8+ T cells. Nat Commun 10, 4987 (2019).

45. Kurachi, M. et al. Chemokine receptor CXCR3 facilitates CD8(+) T cell differentiation into short-lived effector cells leading to memory degeneration. J Exp Med 208, 1605–1620 (2011).

46. Rutella, S. et al. Immune dysfunction signatures predict outcomes and define checkpoint blockade-unresponsive microenvironments in acute myeloid leukemia. J Clin Invest 132, e159579 (2022).

47. Wenk, C. et al. Direct modulation of the bone marrow mesenchymal stromal cell compartment by azacitidine enhances healthy hematopoiesis. Blood Adv 2, 3447–3461 (2018).

48. Platzbecker, U. et al. Efficacy and safety of luspatercept versus epoetin alfa in erythropoiesis-stimulating agent-naive, transfusion-dependent, lower-risk myelodysplastic syndromes (COMMANDS): interim analysis of a phase 3, open-label, randomised controlled trial. Lancet 402, 373–385 (2023).

49. Overacre-Delgoffe, A. E. et al. Interferon-γ Drives Treg Fragility to Promote Anti-tumor Immunity. Cell 169, 1130–1141.e11 (2017).

50. Boukhaled, G. M., Harding, S. & Brooks, D. G. Opposing Roles of Type I Interferons in Cancer Immunity. Annu Rev Pathol 16, 167–198 (2021).

51. Yano, S., Ghosh, P., Kusaba, H., Buchholz, M. & Longo, D. L. Effect of Promoter Methylation on the Regulation of IFN-γ Gene During In Vitro Differentiation of Human Peripheral Blood T Cells into a Th2 Population. The Journal of Immunology 171, 2510–2516 (2003).

52. Chiappinelli, K. B. et al. Inhibiting DNA methylation causes an interferon response in cancer via dsRNA including endogenous retroviruses. Cell 162, 974–986 (2015).

53. Roulois, D. et al. DNA-demethylating agents target colorectal cancer cells by inducing viral mimicry by endogenous transcripts. Cell 162, 961–973 (2015).

54. Heinhuis, K. M. et al. Enhancing antitumor response by combining immune checkpoint inhibitors with chemotherapy in solid tumors. Annals of Oncology 30, 219–235 (2019).

55. Miller, B. C. et al. Subsets of exhausted CD8+ T cells differentially mediate tumor control and respond to checkpoint blockade. Nat Immunol 20, 326–336 (2019).

56. Shallis, R. M. et al. Immune Checkpoint Inhibitor Therapy for Acute Myeloid Leukemia and Higher-Risk Myelodysplastic Syndromes: A Single-Center Experience. Blood 134, 1330 (2019).

57. Khoury, J. D. et al. The 5th edition of the World Health Organization Classification of Haematolymphoid Tumours: Myeloid and Histiocytic/Dendritic Neoplasms. Leukemia 36, 1703–1719 (2022).

58. Greenberg, P. L. et al. Revised International Prognostic Scoring System for Myelodysplastic Syndromes. Blood 120, 2454–2465 (2012).

59. Cheson, B. D. et al. Clinical application and proposal for modification of the International Working Group (IWG) response criteria in myelodysplasia. Blood 108, 419–425 (2006).

60. Döhner, H. et al. Diagnosis and management of AML in adults: 2022 recommendations from an international expert panel on behalf of the ELN. Blood 140, 1345–1377 (2022).

61. Vakrakou, A. G. et al. Specific myeloid signatures in peripheral blood differentiate active and rare clinical phenotypes of multiple sclerosis. Front Immunol 14, 1071623 (2023).

62. Nowicka, M. et al. CyTOF workflow: differential discovery in high-throughput high-dimensional cytometry datasets. F1000Res 6, 748 (2017).

63. Bagwell, C. B. et al. Multi-site reproducibility of a human immunophenotyping assay in whole blood and peripheral blood mononuclear cells preparations using CyTOF technology coupled with Maxpar Pathsetter, an automated data analysis system. Cytometry B Clin Cytom 98, 146–160 (2020).

64. Hao, Y. et al. Integrated analysis of multimodal single-cell data. Cell 184, 3573–3587.e29 (2021).

65. Korsunsky, I. et al. Fast, sensitive and accurate integration of single-cell data with Harmony. Nat Methods 16, 1289–1296 (2019).

66. Kuleshov, M. V. et al. Enrichr: a comprehensive gene set enrichment analysis web server 2016 update. Nucleic Acids Res 44, W90–97 (2016).

67. Xie, Z. et al. Gene Set Knowledge Discovery with Enrichr. Curr Protoc 1, e90 (2021).

68. Cao, J. et al. The single-cell transcriptional landscape of mammalian organogenesis. Nature 566, 496–502 (2019).

69. Huynh-Thu, V. A., Irrthum, A., Wehenkel, L. & Geurts, P. Inferring regulatory networks from expression data using tree-based methods. PLoS One 5, e12776 (2010).

70. Zhu, C. et al. Single-cell transcriptomics dissects hematopoietic cell destruction and T-cell engagement in aplastic anemia. Blood 138, 23–33 (2021).

