## Supplemental Files for "Single-cell analysis of bone marrow CD8+ T cells in Myeloid Neoplasms predicts response to treatment with Azacitidine"

**Supplementary Materials**

Supplementary Figures 1-15

Supplementary Tables 1-4

Data file 1: List of Differentially expressed genes (DEGs) between all clusters used for the characterization of clusters.

Data file 2: List of DEGs for all clusters between MDS and AML samples.

Data file 3: List of DEGs for all clusters between CR and FAIL samples.

**Supplementary Figures**

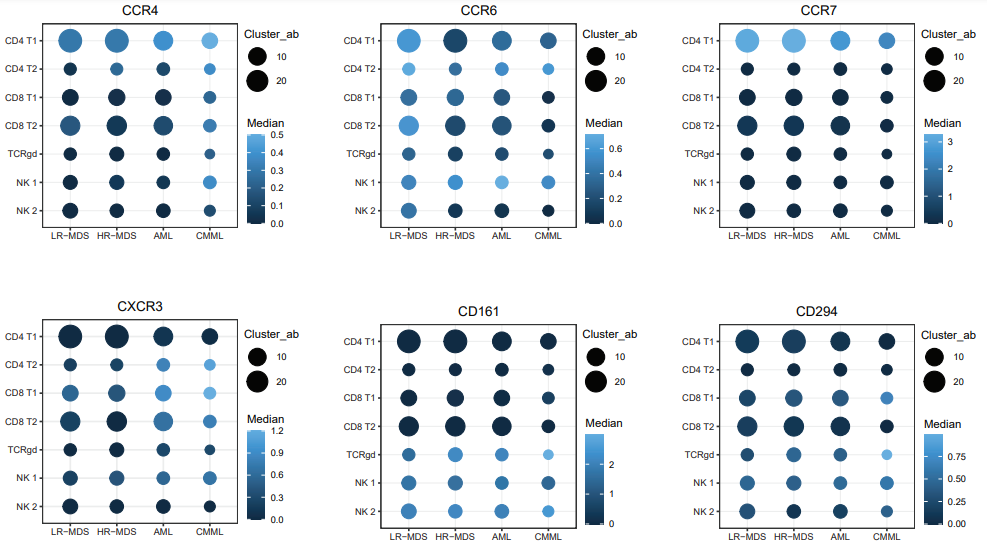

**Supplementary Fig. 1. Expression of cell activation markers in clusters of T and NK cells.** Dot plot displaying the expression level of the activation markers CCR4, CCR6, CCR7, CXCR3, CD161 and CD294 in all T and NK cell clusters. The frequency of each cell cluster is depicted by the dot size, while the color of each dot exhibits the median expression level of each marker for each cell cluster.

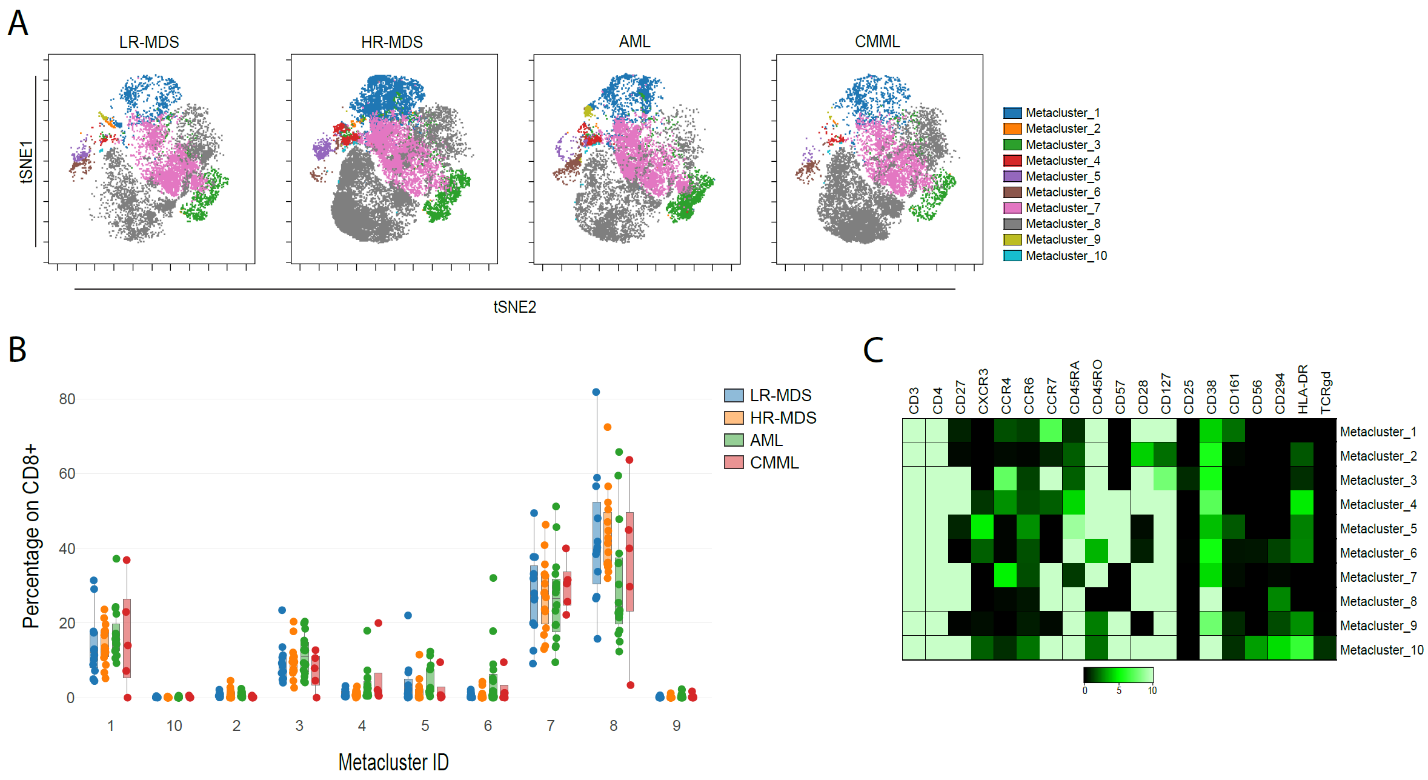
**Supplementary Fig. 2. FlowSOM analysis of bone marrow CD4^+^ T cells from patients with MDS, AML and CMML.** (**A**) Targeted FlowSOM analysis of BM CD4^+^ T cells generated 10 metaclusters, which are projected onto the viSNE plots. viSNE plots from one representative sample of each group are shown here (LR-MDS; n=12, HR-MDS; n=15, AML; n=16, CMML; n=5). (**B**) Boxplots showing the percentage of each cell metacluster expressed as (%) of total CD4^+^ T cells. (**C**) Heatmap depicting the expression level of all T-related markers between the metaclusters. Kruskal Wallis test was used in B.

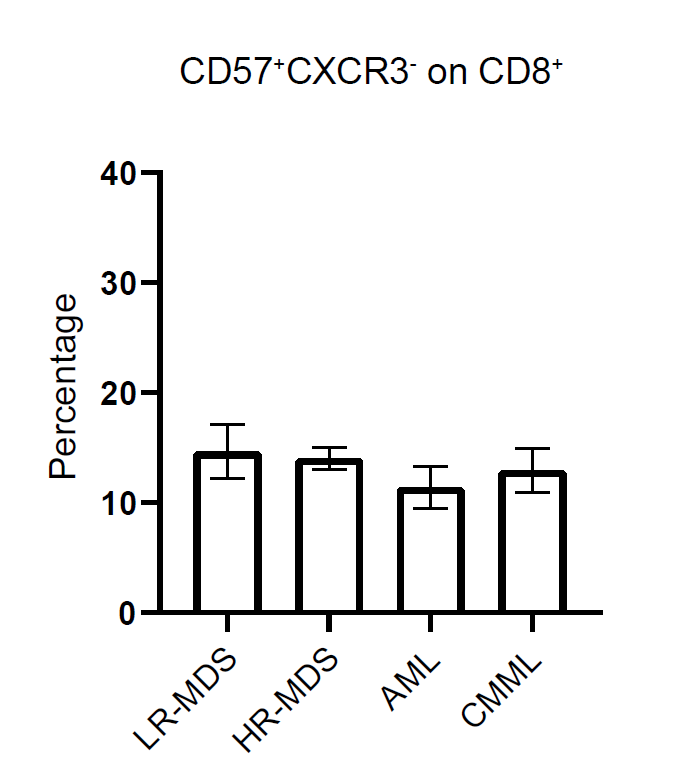

**Supplementary Fig. 3. Frequency of CD57^+^CXCR3^-^ CD8^+^ T cells.** Box plot displaying the cell percentage of the CD57^+^CXCR3^-^ subpopulation on total CD8^+^ T cells (LR-MDS; n=7, HR-MDS; n=27, AML; n=20, CMML; n=10). Data are presented as mean ± SEM. One-way ANOVA followed by "two-stage" Benjamini, Krieger, & Yekutieli multiple comparison test was used.

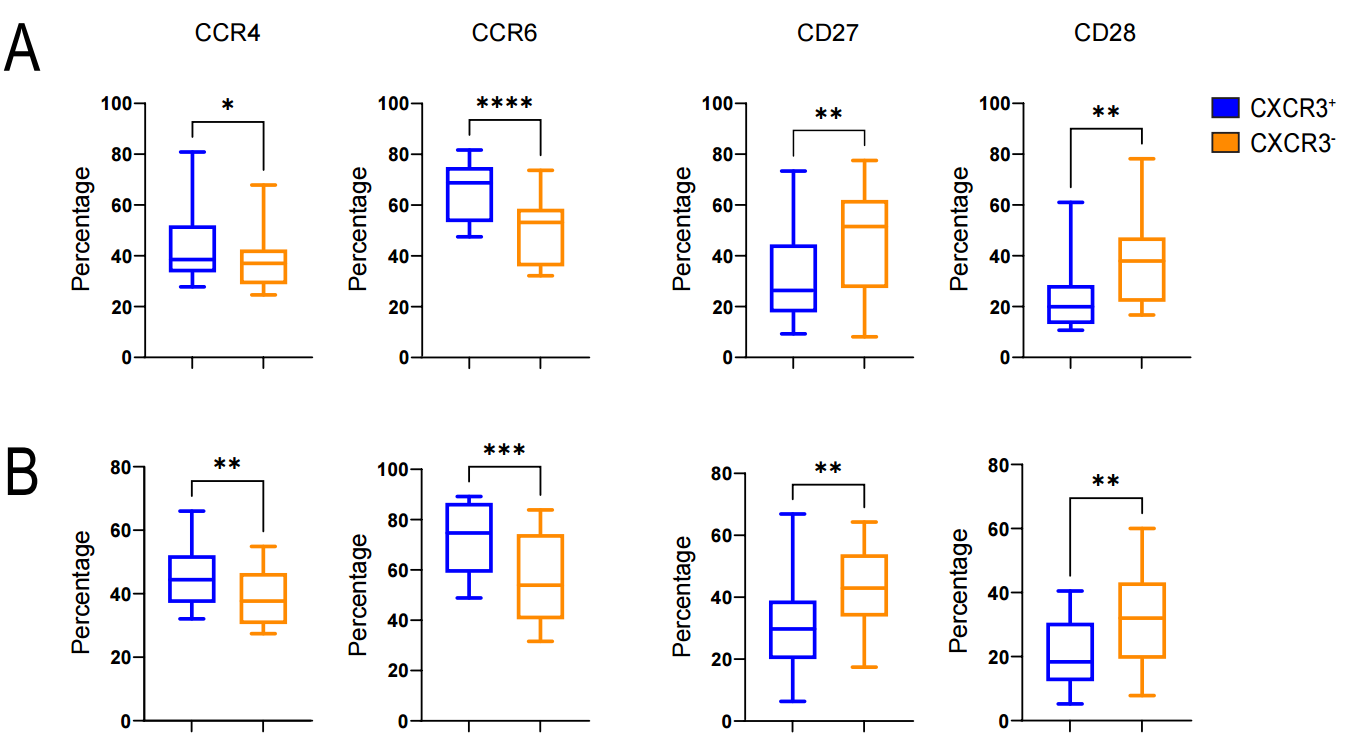

**Supplementary Fig. 4. Immunophenotypic profiling of CD57^+^CXCR3^+^ CD8^+^ T cells by CyTOF.** (**A-Β**) Expression of CD27, CD28, CCR4 and CCR6 in CD8^+^CD57^+^CXCR3^+^ and CD8^+^CD57^+^CXCR3^-^ T cells of HR-MDS (n=15) and AML patients (n=16) respectively. Wilcoxon signed-rank test, *P < 0.05, **P < 0.01, ***P < 0.001, ****P < 0.0001.

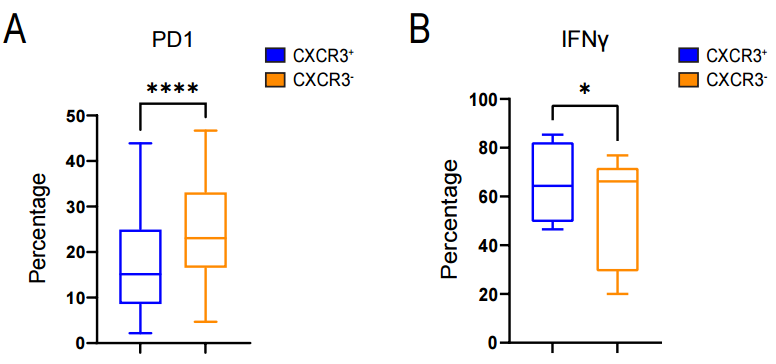

**Supplementary Fig. 5. Immunophenotypic profiling of CD57^+^CXCR3^+^ CD8^+^ T cells by Flow cytometry.** Expression of PD1 on CD8^+^CD57^+^CXCR3^+^ and CD8^+^CD57^+^CXCR3^-^ T cells (n=41, HR-MDS; n=24 and AML; n=17), as assessed by flow cytometry. Wilcoxon signed-rank test, *P < 0.05, **P < 0.01, ***P < 0.001, ****P < 0.0001.

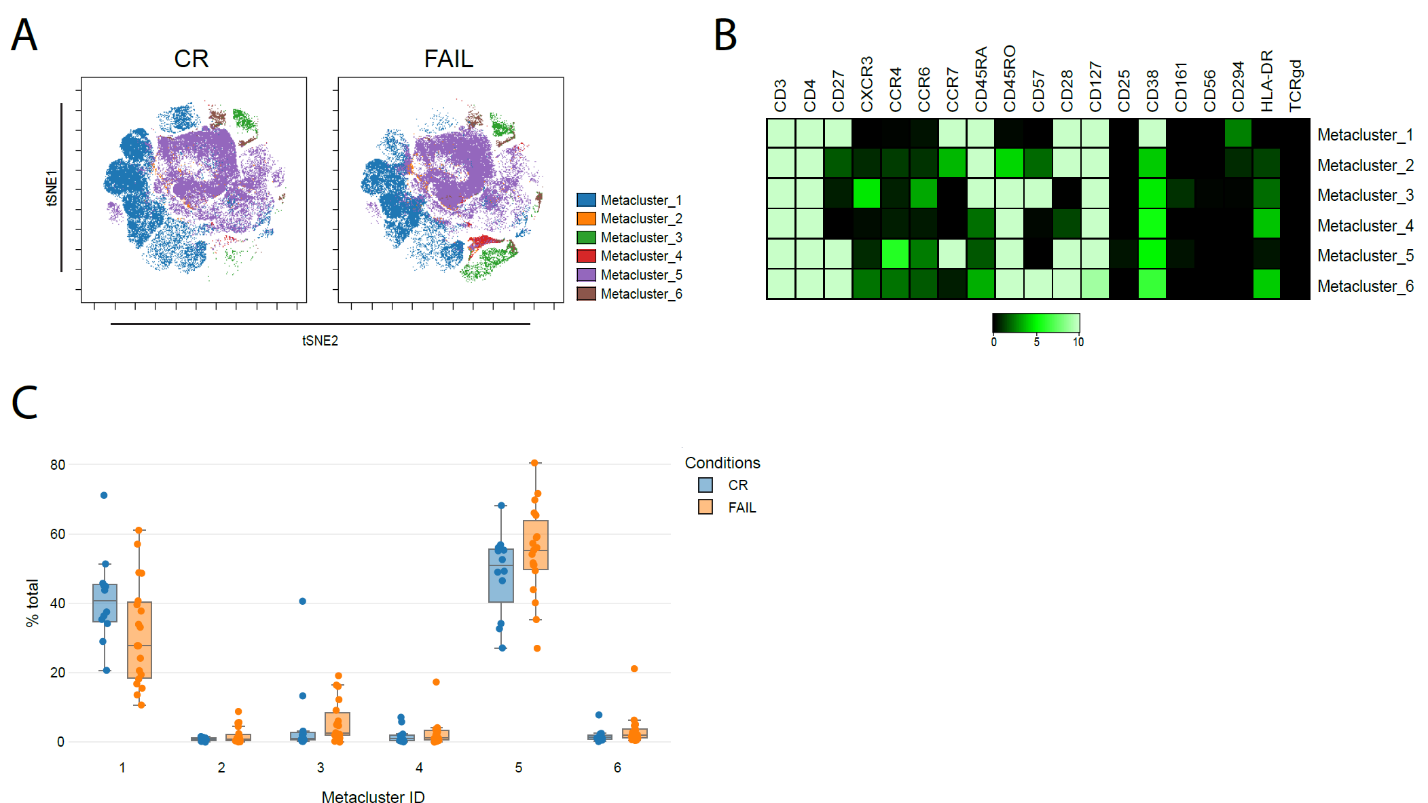
**Supplementary Fig. 6. FlowSOM analysis of BM CD4^+^ T cells in responders and non-responders to AZA.** (**A**) Targeted FlowSOM analysis of BM CD4^+^ T cells generated 6 metaclusters, which are projected onto the viSNE plots. viSNE plots from one representative sample of each group are shown here. (**B**) Boxplots showing the percentage of each cell metacluster expressed as (%) of total CD4^+^ T cells. (**C**) Heatmap depicting the expression level of the cell surface markers. Mann-Whitney U test was used in B.

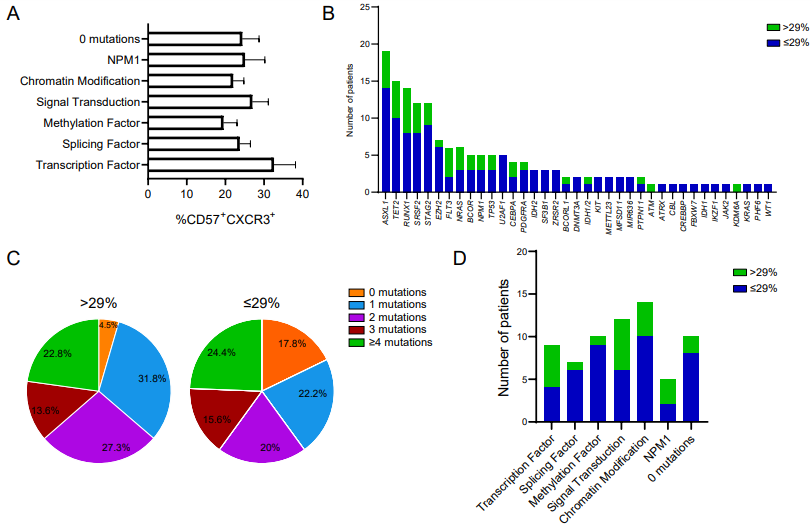

**Supplementary Fig. 7. The presence of specific mutations and their abundance is not linked to the frequency of CD57^+^CXCR3^+^CD8^+^ T cell subset in HR-MDS and AML patients.** (**A**) Frequency of CD57^+^CXCR3^+^CD8^+^ T cells based on oncogenic mutations. (**B**) Proportion of driver mutations in patients with a frequency of >29% (n=22) or ≤29% (n=45) of CD57^+^CXCR3^+^CD8^+^ T cells. (**C**) Number of oncogenic mutations prior to the initiation of treatment with AZA. (**D**) Association between the type of oncogenic mutation and the frequency of CD57^+^CXCR3^+^CD8^+^ T cells.

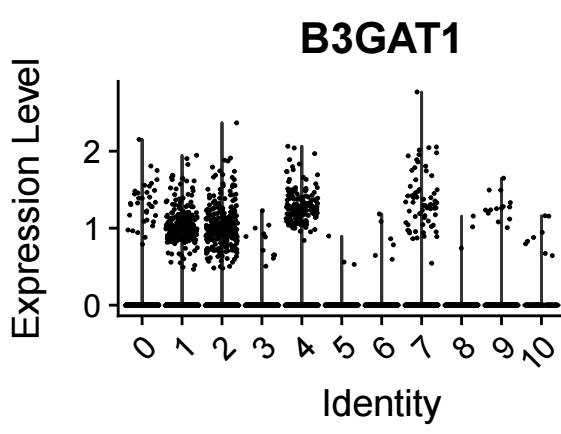

**Supplementary Fig. 8. Clusters 1, 2 and 4 displayed increased expression of the *B3GAT1* gene.** Dot plot showing the differential expression of *B3GAT1* gene between all clusters.

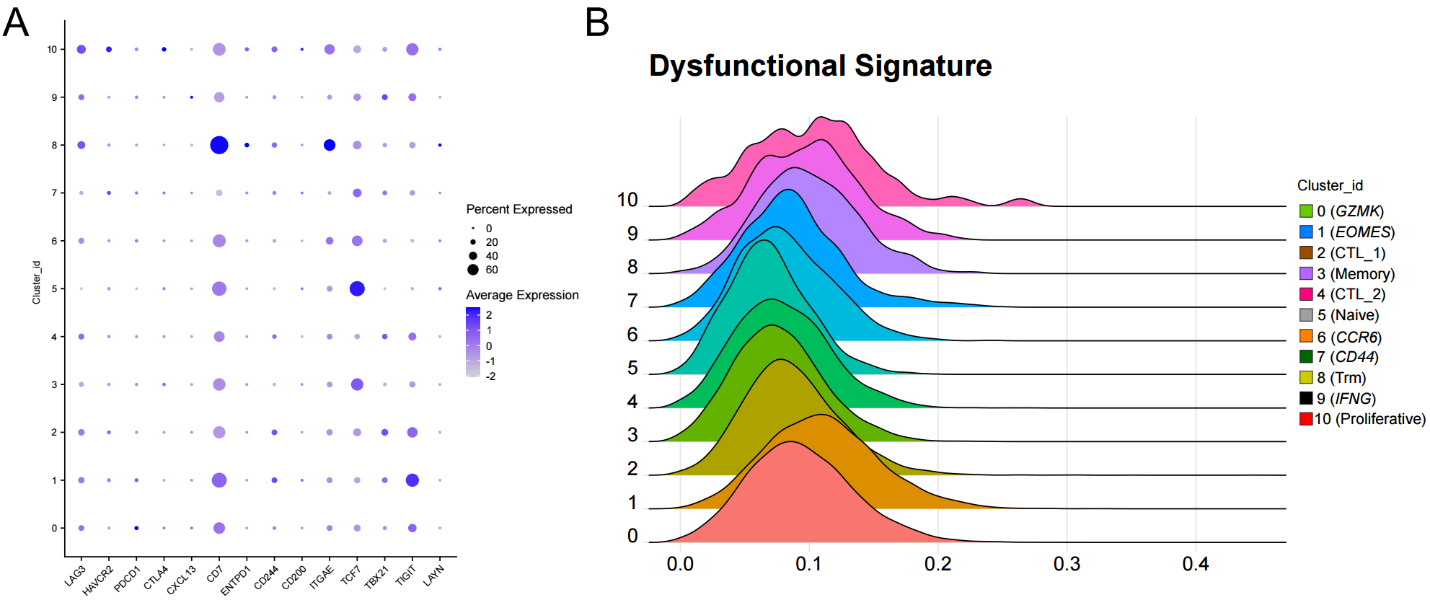
**Supplementary Fig. 9. Characterization of the exhaustion molecular signature between all clusters. (A)** Bubble plot depicting the average expression of representative exhaustion-associated genes used to characterize the clusters. **(B)** Ridgeline plots displaying the dysfunctional/exhaustion signature score for each cell cluster.

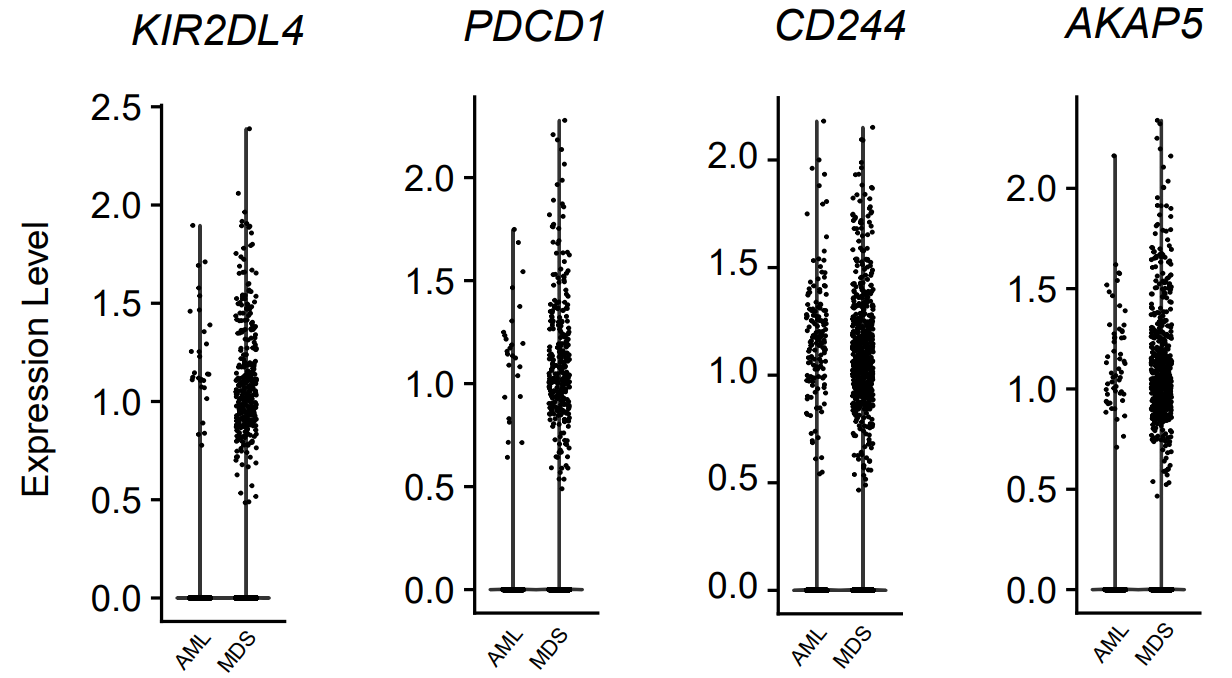

**Supplementary Fig. 10. Dot plots showing the expression level of differentially expressed genes involved in the dysfunctional score of Cluster 1 (*EOMES*) from patients with HR-MDS and secondary AML.**

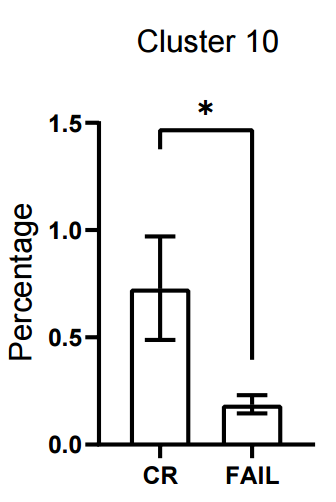

**Supplementary Fig. 11. The frequency of cluster 10 was increased in patients that achieved complete remission.** Boxplot showing the percentage of cluster 10 expressed as (%) of total CD8^+^ T cells (CR; n=4 and FAIL; n=5). Data are presented as mean ± SEM. Unpaired Student’s t test, *P < 0.05.

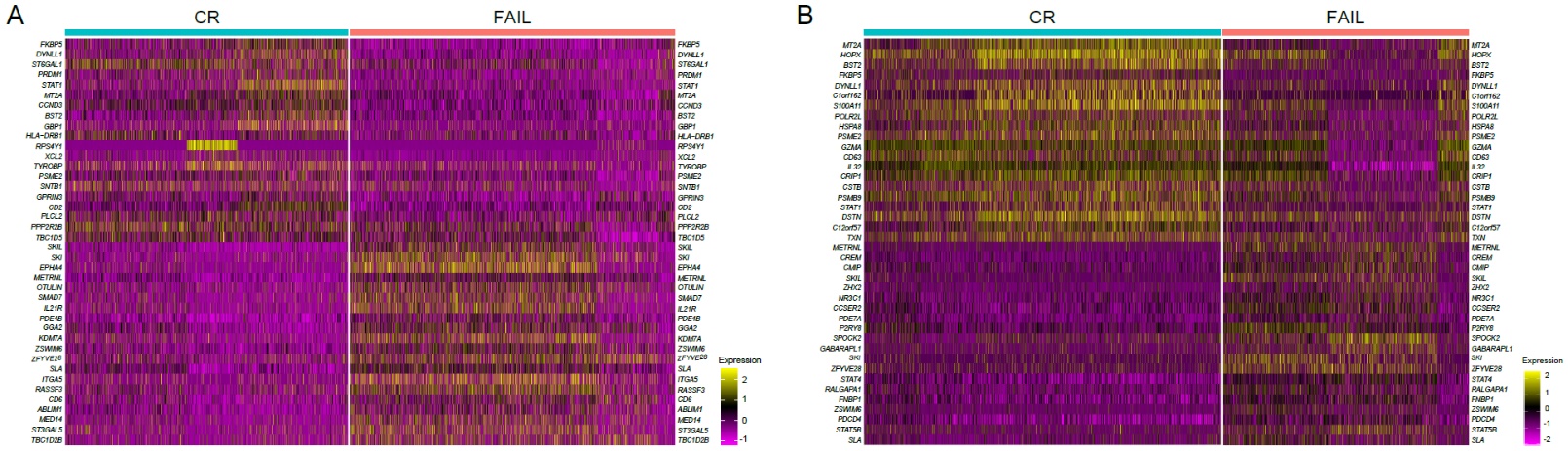

**Supplementary Fig. 12. Top differentially expressed genes (DEGs) of clusters 2 and 4 between CR and FAIL patients**. (**A-B**) Heatmaps displaying the top 20 DEGs of each group in clusters 2 and 4 respectively.

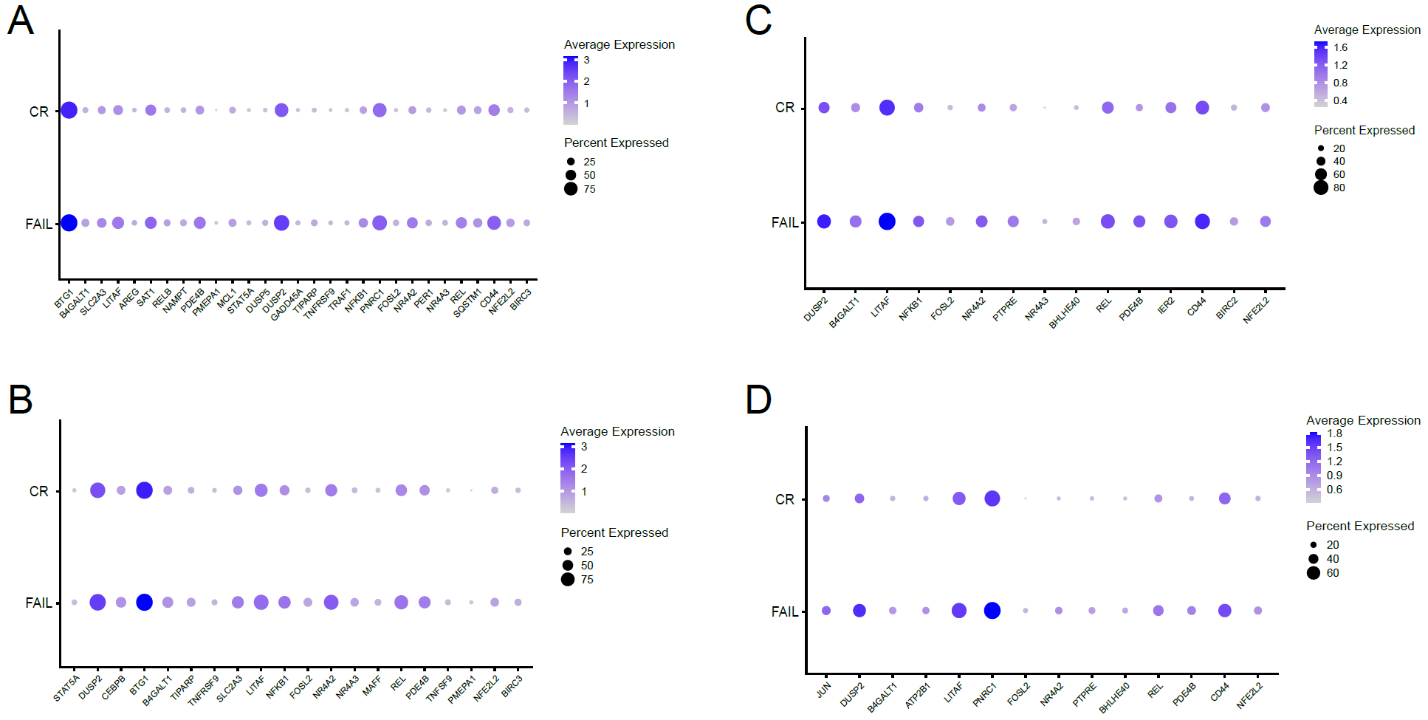

**Supplementary Fig. 13. Bone marrow-derived CD8^+^ T cells of non-responders (FAIL) displayed a positive enrichment of the TNF-α signaling via NF-κΒ, compared to responders (CR), at the scRNA level.** (**A-D**) Dot plots showing the expression level of differentially expressed genes involved in the enrichment of the TNF-α signaling via NF-κΒ pathway, for clusters 0,1,2 and 4 respectively.

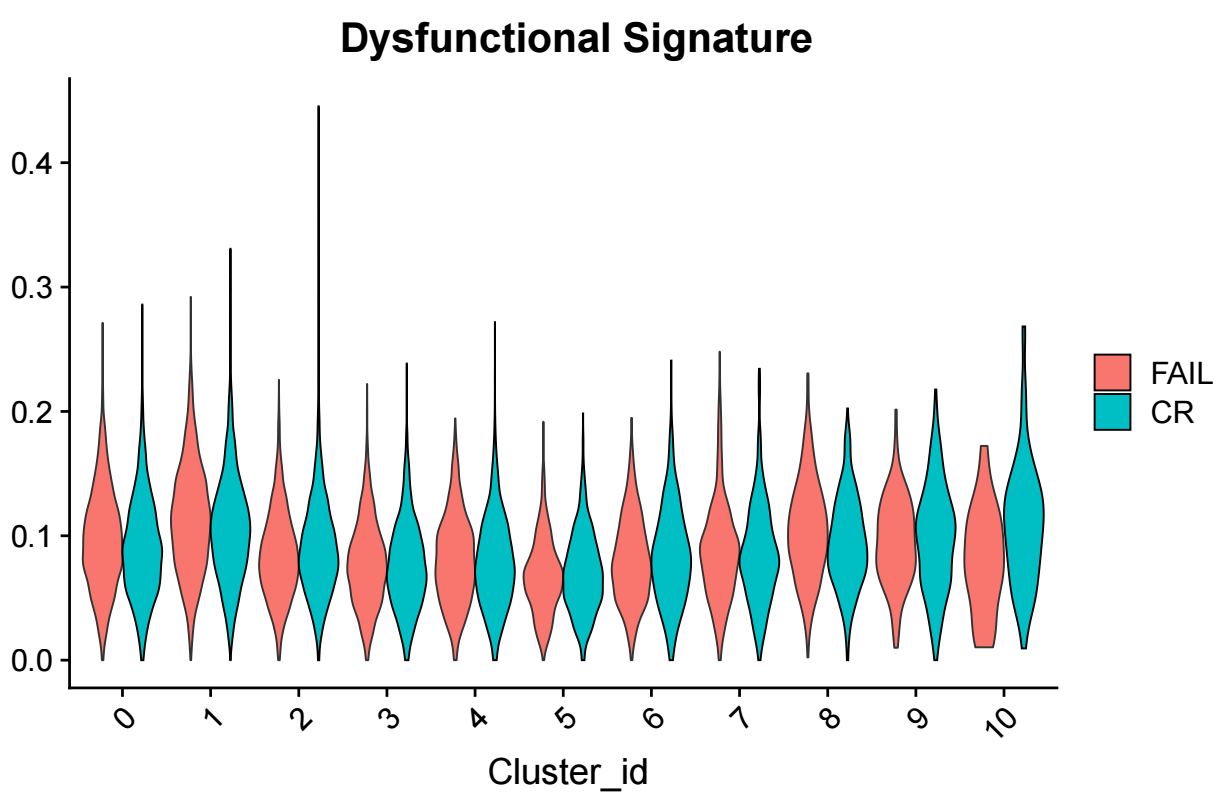

**Supplementary Fig. 14. Outcome of AZA treatment is not associated with a dysfunctional signature of BM CD8^+^ T cells in patients with HR-MDS and secondary AML.** Violin plots depicting the dysfunctional/exhaustion score across all clusters between CR and FAIL patients.

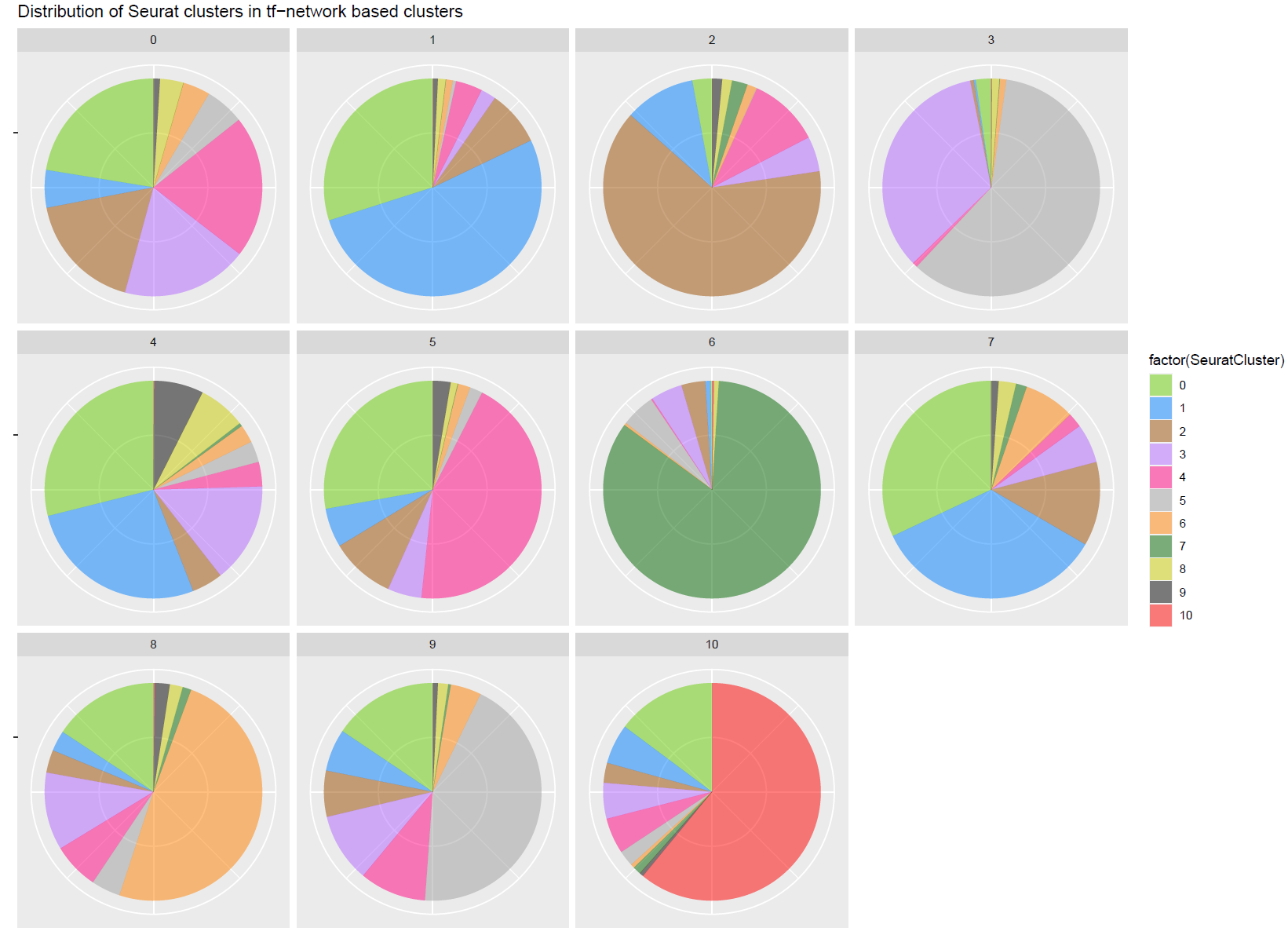
**Supplementary Fig. 15. Distribution of Seurat-defined clusters in TF−network based clusters.** Pie charts illustrating the composition of each regulon cluster in terms of cell types, which were defined based on single-cell gene expression data.

**Supplementary Table 1**. Patients under treatment with azacitidine (n=77); descriptive statistics and uni-/multivariate analysis for response to treatment. OR>1 favors response to treatment.

|  |  | Response to treatment | | | |
| --- | --- | --- | --- | --- | --- |
|  |  | Univariate analysis | | Multivariate analysis | |
| Parameter | Median (range)†, N (%)‡ | OR (±95% CI) | P value | OR (±95% CI) | P value |
| CD57(+)/CXCR3(+)† |  |  |  |  |  |
| *%* | 24.3 (2.9-70.7) | 0.918 (0.875-0.964) | **0.001** |  |  |
| CD57(+)/CXCR3(+)‡ |  |  |  |  |  |
| *≤29%* | 50 (64.9) | 1.000 | **0.001** | 1.000 | **0.006** |
| *>29%* | 27 (35.1) | 0.138 (0.045-0.424) |  | 0.151 (0.039-0.584) |  |
| WHO classification |  |  |  |  |  |
| *HR-MDS* | 37 (48.1) | 1.000 |  | 1.000 |  |
| *AML* | 29 (37.7) | 0.357 (0.130-0.978) | **0.045** | 0.419 (0.059-2.958) | 0.383 |
| *CMML* | 11 (14.3) | 0.625 (0.160-2.441) | 0.499 | 1.364 (0.166-11.223) | 0.773 |
| Sex |  |  |  |  |  |
| *Females* | 23 (29.9) | 1.000 | 0.820 | 1.000 | 0.314 |
| *Males* | 54 (70.1) | 1.121 (0.419-2.994) |  | 1.938 (0.534-7.031) |  |
| Age |  |  |  |  |  |
| *years* | 72.2 (48.6-93.6) | 1.028 (0.978-1.081) | 0.281 | 1.025 (0.960-1.094) | 0.466 |
| BM blasts |  |  |  |  |  |
| *%* | 16.0 (5.0-92.0) | 0.987 (0.967-1.006) | 0.181 | 1.004 (0.968-1.041) | 0.834 |
| Hb |  |  |  |  |  |
| *g/dL* | 9.1 (5.4-13.9) | 0.950 (0.675-1.336) | 0.766 | 1.069 (0.701-1.631) | 0.757 |
| ANC |  |  |  |  |  |
| */10^-9^ L* | 1.6 (0.0-49.1) | 0.984 (0.923-1.048) | 0.612 | 0.943 (0.860-1.033) | 0.208 |
| PLT |  |  |  |  |  |
| */10^-9^ L* | 64.0 (5.0-422.0) | 1.000 (0.995-1.005) | 0.873 | 0.998 (0.991-1.005) | 0.581 |
| Cytogenetics (IPSS-R) |  |  |  |  |  |
| *Very Good* | 1 (1.4) | 1.000 | 0.316 | 1.000 | 0.262 |
| *Good* | 40 (55.6) |  |  |  |  |
| *Intermediate* | 20 (27.8) | 0.541 (0.208-1.412) |  | 0.512 (0.159-1.649) |  |
| *Poor* | 2 (2.8) |  |  |  |  |
| *Very Poor* | 9 (12.5) |  |  |  |  |
| IPSS-R score |  |  |  |  |  |
|  | 6.0 (4.0-9.5) | 1.005 (0.704-1.434) | 0.979 |  |  |
| Total mutations |  |  |  |  |  |
|  | 2.0 (0-14) | 1.117 (0.916-1.361) | 0.275 | 1.312 (0.969-1.777) | 0.079 |
| Mutation Category |  |  |  |  |  |
| *Chromatin Modification* | 13 (16.9) | 0.473 (0.132-1.694) | 0.250 |  |  |
| *Transcription Factor* | 10 (13.0) | 1.531 (0.425-5.520) | 0.515 |  |  |
| *Methylation Factor* | 12 (15.6) | 1.531 (0.425-5.520) | 0.515 |  |  |
| *Signal Transduction* | 14 (18.2) | 1.034 (0.313-3.422) | 0.956 |  |  |
| *Splicing Factor* | 7 (9.1) | 0.694 (0.154-3.133) | 0.694 |  |  |
| *NPM1* | 9 (11.7) | 1.583 (0.391-6.416) | 0.520 |  |  |
| Mutation |  |  |  |  |  |
| *Any* | 64 (83.1) | 0.967 (0.292-3.198) | 0.956 |  |  |
| *ASXL1* | 18 (23.4) | 0.625 (0.215-1.813) | 0.387 |  |  |
| *NRAS* | 8 (10.4) | 0.563 (0.130-2.435) | 0.442 |  |  |
| *RUNX1* | 16 (20.8) | 0.470 (0.146-1.514) | 0.206 |  |  |
| *SRSF2* | 12 (15.6) | 0.708 (0.209-2.400) | 0.580 |  |  |
| *STAG2* | 9 (11.7) | 1.531 (0.425-5.520) | 0.424 |  |  |
| *TET2* | 19 (24.7) | 0.948 (0.328-2.741) | 0.922 |  |  |
| IPSS-M score |  |  |  |  |  |
|  | 2.0 (1.0-4.0) | 0.913 (0.613-1.360) | 0.653 |  |  |
| Response |  |  |  |  |  |
| *CR* | 35 (45.5) |  |  |  |  |
| *Fail* | 42 (54.5) |  |  |  |  |
| Overall Survival |  |  |  |  |  |
| *months* | 18.5 (13.8-23.2) |  |  |  |  |
| Follow-up |  |  |  |  |  |
| *months* | 87.3 (76.9-97.7) |  |  |  |  |

† continuous variables; ‡ discrete variables

**Supplementary Table 2.** Details of patient samples used in scRNA-seq.

| **Sample Title** | **Phenotype** | **(%) Blasts** | **Response** | **Age (years)** |
| --- | --- | --- | --- | --- |
| MDS1 | HR-MDS | 8 | CR | 73 |
| MDS2 | HR-MDS | 16 | CR | 52 |
| MDS3 | HR-MDS | 8 | FAIL | 69 |
| MDS4 | HR-MDS | 18 | FAIL | 79 |
| AML1 | Secondary AML | 65 | CR | 72 |
| AML2 | Secondary AML | 60 | CR | 72 |
| AML3 | Secondary AML | 25 | FAIL | 76 |
| AML4 | Secondary AML | 55 | FAIL | 53 |
| AML5 | Secondary AML | 77 | FAIL | 80 |

**Supplementary Table 3.** Antibodies used for mass cytometry

| **Target** | **Clone** | **Metal** | **Localization** | **Supplier** |
| --- | --- | --- | --- | --- |
| CD3 | UCHT1 | 170 Er | Cell-surface | Standard Biotools (MDIPA backbone SB) |
| CD4 | RPA‐T4 | 145 Nd | Cell-surface | Standard Biotools (MDIPA backbone SB) |
| CD8 | RPA‐T8 | 146 Nd | Cell-surface | Standard Biotools (MDIPA backbone SB) |
| CD11c | Bu15 | 147 Sm | Cell-surface | Standard Biotools (MDIPA backbone SB) |
| CD14 | 63D3 | 168 Er | Cell-surface | Standard Biotools (MDIPA backbone SB) |
| CD16 | 3G8 | 148 Nd | Cell-surface | Standard Biotools (MDIPA backbone SB) |
| CD19 | HIB19 | 144 Nd | Cell-surface | Standard Biotools (MDIPA backbone SB) |
| CD20 | 2H7 | 171 Yb | Cell-surface | Standard Biotools (MDIPA backbone SB) |
| CD25 | BC96 | 153 Eu | Cell-surface | Standard Biotools (MDIPA backbone SB) |
| CD27 | O323 | 154 Sm | Cell-surface | Standard Biotools (MDIPA backbone SB) |
| CD28 | CD28.2 | 160 Gd | Cell-surface | Standard Biotools (MDIPA backbone SB) |
| CD38 | HB‐7 | 161 Dy | Cell-surface | Standard Biotools (MDIPA backbone SB) |
| CD45 | HI30 | 89 Y | Cell-surface | Standard Biotools (MDIPA backbone SB) |
| CD45RA | HI100 | 150 Nd | Cell-surface | Standard Biotools (MDIPA backbone SB) |
| CD45RO | UCHL1 | 149 Sm | Cell-surface | Standard Biotools (MDIPA backbone SB) |
| CD56 | NCAM16.2 | 163 Dy | Cell-surface | Standard Biotools (MDIPA backbone SB) |
| CD57 | HCD57 | 155 Gd | Cell-surface | Standard Biotools (MDIPA backbone SB) |
| CD66b | G10F5 | 172 Yb | Cell-surface | Standard Biotools (MDIPA backbone SB) |
| CD123 (IL3RA) | 6H6 | 143 Nd | Cell-surface | Standard Biotools (MDIPA backbone SB) |
| CD127 | A019D5 | 176 Yb | Cell-surface | Standard Biotools (MDIPA backbone SB) |
| CD161 | HP‐3G10 | 151 Eu | Cell-surface | Standard Biotools (MDIPA backbone SB) |
| CD294 | BM16 | 166 Er | Cell-surface | Standard Biotools (MDIPA backbone SB) |
| CCR4 | L291H4 | 152 Sm | Cell-surface | Standard Biotools (MDIPA backbone SB) |
| CCR6 | G034E3 | 141 Pr | Cell-surface | Standard Biotools (MDIPA backbone SB) |
| CCR7 | G043H7 | 167 Er | Cell-surface | Standard Biotools (MDIPA backbone SB) |
| CXCR3 (CD183) | G025H7 | 156 Gd | Cell-surface | Standard Biotools (MDIPA backbone SB) |
| CXCR5 (CD185) | J252D4 | 158 Gd | Cell-surface | Standard Biotools (MDIPA backbone SB) |
| HLA-Dr | LN3 | 173 Yb | Cell-surface | Standard Biotools (MDIPA backbone SB) |
| IgD | IA6‐2 | 174 Yb | Cell-surface | Standard Biotools (MDIPA backbone SB) |
| TCRγδ | B1 | 164 Dy | Cell-surface | Standard Biotools (MDIPA backbone SB) |

**Supplementary Table 4.** Antibodies used for conventional flow cytometry staining

| **Target** | **Clone** | **Fluorochrome** | **Localization** | **Supplier** | **Cat No.** |
| --- | --- | --- | --- | --- | --- |
| CD3 | SK7 | BV421 | Cell-surface | Biolegend | 344834 |
| CD4 | SK3 | APC/Cy7 | Cell-surface | Biolegend | 344616 |
| CD8 | SK1 | BV510 | Cell-surface | Biolegend | 344732 |
| CXCR3 (CD183) | G025H7 | APC | Cell-surface | Biolegend | 353708 |
| CD279 (PD1) | EH12.2H7 | PE/Cy7 | Cell-surface | Biolegend | 329918 |
| CD57 | NK-1 | FITC | Cell-surface | BD Biosciences | 555619 |
